# Genome-wide association study identifies new loci associated with OCD

**DOI:** 10.1101/2024.03.06.24303776

**Authors:** Nora I Strom, Matthew W Halvorsen, Chao Tian, Christian Rück, Gerd Kvale, Bjarne Hansen, Jonas Bybjerg-Grauholm, Jakob Grove, Julia Boberg, Judith Becker Nissen, Thomas Damm Als, Thomas Werge, Elles de Schipper, Bengt Fundin, Christina Hultman, Kira D. Höffler, Nancy Pedersen, Sven Sandin, Cynthia Bulik, Mikael Landén, Elinor Karlsson, Kristen Hagen, Kerstin Lindblad-Toh, Nordic OCD and Related Disorders Consortium (NORDiC), 23andMe Research Team, PGC TS/OCD working group, David M. Hougaard, Sandra M. Meier, Stéphanie Le Hellard, Ole Mors, Anders D. Børglum, Jan Haavik, David A. Hinds, David Mataix-Cols, James J Crowley, Manuel Mattheisen

## Abstract

To date, four genome-wide association studies (GWAS) of obsessive-compulsive disorder (OCD) have been published, reporting a high single-nucleotide polymorphism (SNP)-heritability of 28% but finding only one significant SNP. A substantial increase in sample size will likely lead to further identification of SNPs, genes, and biological pathways mediating the susceptibility to OCD. We conducted a GWAS meta-analysis with a 2-3-fold increase in case sample size (OCD cases: N = 37,015, controls: N = 948,616) compared to the last OCD GWAS, including six previously published cohorts (OCGAS, IOCDF-GC, IOCDF-GC-trio, NORDiC-nor, NORDiC-swe, and iPSYCH) and unpublished self-report data from 23andMe Inc. We explored the genetic architecture of OCD by conducting gene-based tests, tissue and celltype enrichment analyses, and estimating heritability and genetic correlations with 74 phenotypes. To examine a potential heterogeneity in our data, we conducted multivariable GWASs with MTAG. We found support for 15 independent genome-wide significant loci (14 new) and 79 protein-coding genes. Tissue enrichment analyses implicate multiple cortical regions, the amygdala, and hypothalamus, while cell type analyses yielded 12 cell types linked to OCD (all neurons). The SNP-based heritability of OCD was estimated to be 0.08. Using MTAG we found evidence for specific genetic underpinnings characteristic of different cohort-ascertainment and identified additional significant SNPs. OCD was genetically correlated with 40 disorders or traits-positively with all psychiatric disorders and negatively with BMI, age at first birth and multiple autoimmune diseases. The GWAS meta-analysis identified several biologically informative genes as important contributors to the aetiology of OCD. Overall, we have begun laying the groundwork through which the biology of OCD will be understood and described.

## Introduction

Obsessive-compulsive disorder (OCD) is a neuropsychiatric disorder characterized by recurrent, unwanted thoughts and/or repetitive behaviors ^1;2^. It is relatively common (1-3% prevalence ^3^, often begins in childhood and although medication and behavioral therapy are useful, symptom control is imperfect, and the course is often chronic. Most individuals with OCD also have a comorbid psychiatric disorder (e.g., tic disorders, anorexia nervosa, mood-, and/or anxiety disorders) ^3;4^ and early onset and tic-related OCD may be etiologically meaningful subtypes of the disorder ^5;6^. OCD is responsible for profound personal and societal costs ^7^ and patients are at a substantial risk of suicide (∼10 times higher than the population prevalence) ^8^ and show an increase in general mortality ^9^. OCD is highly heritable (∼50%) ^10;11^, and first-degree relatives of affected individuals have a 4-8 times increased risk of developing the disorder ^11–13^. It is hoped that the identification of specific genes and biological pathways mediating susceptibility to OCD will lead to improved understanding and perhaps to improved detection, differential diagnosis, and/or treatment of OCD. Unfortunately, several OCD linkage studies (reviewed by Pauls et al. ^14^) and >100 candidate gene studies (meta-analyzed by Taylor et al. ^15^) have produced inconsistent results. More recently, there have been four published genome-wide association studies (GWAS) of OCD. The first ^16^ included 1,465 cases, 5,557 controls, and 400 trios. No genome-wide significant loci were identified, but polygenic risk analysis revealed overlap with Tourette syndrome and indicated that increased sample size will likely reveal significant loci ^17^. A second OCD GWAS ^18^ had 1,406 cases and 3,655 controls and although a third GWAS meta-analysis ^19^ of these two studies revealed substantial heritability of OCD risk based on common variation (0.28 *±* 0.04), it failed to identify genome-wide significant loci. The fourth GWAS, recently published on medRxiv ^20^, was comprised of a total of 14,140 OCD cases and 562,117 controls and revealed the first genome-wide significant hit for OCD on chromosome 3. Sample size has clearly been the limiting factor for gene discovery in OCD. Here we present GWAS results obtained from a sample with a ∼ 2-3 fold increase in case sample size (37,015 cases and 948,616 controls) over the recent medRxiv paper. Cases and controls were derived from 1) a previous OCD GWAS meta-analysis mentioned above ^19^, 2) ongoing studies in Scandinavia (NORDiC ^21^ and the Danish iPSYCH cohort ^22;23^), which were also included in the most recent OCD GWAS meta-analysis ^20^, and 3) unpublished self-report OCD cases and controls from 23andMe Inc.

## Methods

### Samples

We analysed genomic data from seven OCD case-control cohorts of European ancestry. The meta-analysis included 1) partially published data from Scandinavia including NORDiC-NOR (365 OCD cases, 315 controls), NORDiC-SWE (977 OCD cases, 3,200 controls ^21^), and iPSYCH (2,678 OCD cases, 10,410 controls) (data was included in the recently published OCD GWAS ^20^; 2) the three datasets from the previously published OCD PGC-freeze 1^19^), comprised of OCGAS ^18^ (986 OCD cases, 1,023 controls; with changed control-inclusion compared to the original publication), IOCDF-GC (1,519 OCD cases, 3,541 controls ^16^), and IOCDF-GC-trio (323 complete trios); and 3) unpublished self-report OCD cases and controls from 23andMe (30,167 OCD cases, 929,804 controls). Together, the GWAS meta-analysis included 37,015 OCD cases and 948,616 controls of European ancestry. We further conducted three separate GWAS analyses, a) combining all clinical cases primarily ascertained for OCD (OCD PGC-freeze1 and NORDiC samples (NORDiC-NOR and NORDiC-SWE), in the following referred to as “freeze1+NORDiC”), b) clinical cases obtained through a population-based sample primarily ascertained for another comorbid psychiatric disorder (iPSYCH), and c) self-report samples (23andMe). We also conducted three multivariable GWAS analyses with MTAG for the same sub-groups as described above. Cohort specific sample and analytic details can be found in the following and Supplementary Table S1 provides an overview of the individual cohorts. Data collections were approved by the relevant institutional review boards at all participating sites. Where required, all participants provided written informed consent.

### GWAS analysis NORDiC-NOR & NORDiC-SWE

#### pre-GWAS Quality Control (QC)

We assembled two separate NORDiC case/control datasets from genotype array data for our GWAS. The first consisted of 647,335 variant calls across a total of 1,107 NORDiC-SWE case and 3,500 Swedish control samples. The second consisted of 482 NORDiC-NOR case and 343 Norwegian-ancestry control samples. The Swedish GWAS was a merger across a total of four different cohorts (one case-only, three control-only) whereas the Norwegian GWAS was a merger across 3 cohorts (two case-only, one control-only). Variant-level QC on the data removed SNP markers where there was evidence of excess missingness in any of the input cohorts, case/control missingness difference, or evidence of clustering artifact. Sample-level QC on the data removed individuals with excess missingness and instances of cryptic relatedness. Sample-level QC also identified a subset of samples of likely European ancestry based on clustering with 1000 genomes data ^24^, ensuring the construction of datasets that are homogeneous and less prone to potential effects of population stratification. The process left us with 492,554 QC-passing variants across the NORDiC-SWE GWAS data (1,107 cases and 3,500 controls) and 479,358 QC-passing variants across the NORDiC-NOR data (407 cases and 340 controls). Please refer to the section ‘GWAS details for NORDiC-NOR & NORDiC-SWE’ in the supplemental text for details specific to the Swedish and Norwegian QC and GWAS analyses.

#### QC and imputation implemented in RICOPILI

We used the Rapid Imputation and COmputational PIpeLIne for Genome-Wide Association Studies (RICOPILI; ^25^) to run an automated round of pre-imputation QC on the genotype data from 492,554 QC-passing variants across the full unpruned datasets of NORDiC-SWE (1,107 cases and 3,500 controls) and 479,358 QC-passing variants across NORDIC-NOR (407 cases and 340 controls), and conducted imputation on the genotype data using the Haplotype Reference Consortium (HRC) reference panel. All subsets of the analysis were done using RICOPILI v2018_Dec_7.001. The pre-imputation RICOPILI QC step involved a series of hard filters on variant and sample level data, including removing variants with pre-sample pruning call rate < 0.95, samples with call rate < 0.98, FHET outside of +/− 0.20, samples with discrepancies between reported and derived sex, and post sample-pruning variants that met any of the following: 1) call rate < 0.98, 2) missing difference > 0.02, 3) invariant positions, 4) minor allele frequency (MAF) > 0.01, 5) Hardy-Weinberg equilibrium (HWE) p < 1×10^*−*6^ in controls, and 6) HWE p < 1×10^*−*10^ in cases. After the QC described we were left with 481,323 QC-passing variants across 1,077 cases and 3,486 controls in NORDiC-SWE, and 469,618 QC-passing variants across 404 cases and 340 controls in NORDiC-NOR.

After sample and variant pruning we next used RICOPILI’s impute_dirsub module to perform imputation using HRC genotypes as a reference panel. In our imputation run we used eagle v2.3.5 for pre-phasing, and minimac3 v2.0.1 for imputation. We derived 3 different imputed callsets from this process: 1) a set of high confidence imputed genotypes (2,771,425 in NORDiC-SWE, 3,043,464 in NORDiC-NOR), 2) a set of imputed best-guess genotypes with medium level accuracy (7,112,906 in NORDiCSWE, 7,277,174 in NORDiC-NOR), and 3) a set of variants for which imputation accuracy was lowered in order to increase the total number of variants included in the imputation (9,021,638 in NORDiC-SWE, 8,964,589 in NORDiC-NOR).

#### GWAS

We ran our GWAS across the largest dataset of imputed variants that were generated during the imputation process on a subset of samples that were of European ancestry. We intersected the RICOPILI QC-pruned set of samples with our own set of relatedness-pruned European ancestry samples described earlier to obtain a set of 977 cases and 3,200 controls for NORDiC-SWE GWAS, and 365 cases and 315 controls for NORDiC-NOR OCD GWAS. We conducted principle component analysis (PCA) on these samples across high-confidence imputed genotypes using the pacer_sub RICOPILI module and tested the first 20 PCs for significant association with sample case/control status (significant = p-value < 0.05/20). We identified PCs 1, 3 and 14 as significant predictors and used them as covariates in the NORDiC-SWE GWAS. Along the same lines we identified PCs 1, 2, 3 and 4 as significant predictors and used them as covariates in the NORDiC-NOR GWAS.

### GWAS analysis iPSYCH

Danish nation-wide population-based case-control samples were collected in the scope of the ‘Danish OCD and Tourette Study’ (DOTS) within ‘The Lundbeck Foundation Initiative for Integrative Psychiatric Research’ (iPSYCH). Individuals were not primarily ascertained for OCD; OCD cases were drawn from cases that also presented a diagnosis of another psychiatric disorder and from controls with a diagnosis of OCD.

Heel prick blood samples had been collected from all babies in Denmark born between 1981 and 2005. Genetic information was obtained by the Statens Serum Institut (SSI) at the Danish Neonatal Screening Biobank (DNSB). The genetic information was linked with the Danish Civil Registration System and thereby coupled with the Danish Psychiatric Central Research Register which collects patient data of individuals treated in psychiatric hospitals or in outpatient psychiatric clinics. OCD cases met ICD10 (F42) criteria, controls were randomly selected from the same birth cohorts and excluded individuals with a F42 diagnosis.

Genotyping was performed on the PsychChip v 1.0 array (Illumina, San Diego, CA, USA) at the Broad Institute of MIT and Harvard (Cambridge, MA, USA). Genotyping and data analysis was performed in 25 batches. Genotype data was processed using RICOPILI performing stringent QC. Samples with a call rate below 95%, with a sex mismatch, between the sex obtained from genotype data and from the register data, as well as related individuals were removed. Principle component analysis was used to exclude ancestral outliers from non-European descent. The data was imputed using the HRC reference panel. The final data set included 2,678 OCD cases and 10,410 controls.

The study was approved by the Regional Scientific Ethics Committee in Denmark and the Danish Data Protection Agency. All analyses of the samples were performed on the secured national GenomeDK high performance-computing cluster in Denmark (https://genome.au.dk). Details of the cohort, data QC and GWAS analysis have been described else-where ^22;26^.

### GWAS analysis OCD PGC-freeze1

The OCD PGC-freeze1 cohort refers to the OCD GWAS meta-analysis previously published by the PGC, comprising data from the ‘OCD Foundation-Genetics Consortium’ (IOCDF-GC) and ‘The OCD Collaborative Genetics Association Study’ (OCGAS). Details of the cohorts and analysis can be found in the primary publications ^16;18;27^. Here, we used the OCGAS and IOCDF-GC data from a recent meta-analysis for which the data was re-analyzed with newly matched controls ^20^.

The IOCDF-GC cohort consists of a case-control sample and a trio sample. The case-control cohort consists of 1,519 European OCD cases and 3,541 matched controls. Part of the controls were drawn from previously genotyped cohorts including the Alzheimer’s Disease Genetics Initiative ^28^, the Center for Applied Genomics (CAG) at Children’s Hospital of Philadelphia (CHOP) ^29^, and the Breast and Prostate Cancer Cohort Consortium (BPC3) ^30^. The trio sample consists of 323 complete trios of European ancestry. Cases and trios were predominantly recruited from OCD specialty clinics. Controls were recruited from Bonn, Germany and from Capetown, South Africa. All cases met DSM-IV diagnosis of OCD, while the controls from Bonn had no lifetime history of any axis I disorders and the controls from Cape Town were unscreened.

Genotyping was performed on the Illumina Human610-Quadv1_B array (Illumina, San Diego, CA, USA). Standard QC was performed using PLINK2^31^. Samples with a call rate *<*98%, sex discrepancy or ambiguous genomic sex, and related samples (pihat*>*0.2) were removed. SNPs with a genotyping rate *<*98%, with a MAF *<* .01, monomorphic SNPs, CNV-targeted SNP probes, strand-ambiguous SNPs with significant allele frequency differences or aberrant LD correlations with neighbouring SNPs based on the entire HapMap2 reference panel, SNPs with differential missing rate between cases and controls (*>*0.02), SNPs with *P<*1×10^*−*6^ (controls) or *P<*1×10^*−*10^ (cases) in HWE, and SNPs with batch effect (P*<*1×10^*−*5^) in control cohorts were removed. The same QC was performed on the trios, additionally removing SNPs with Mendelian errors. For each trio, the transmitted and untransmitted alleles were converted into one case and one pseudo-control. Multidimensional scaling analysis (MDS) was performed and samples with significant outliers in the first five dimensions or when there were no matching cases and controls for each dimension, were removed. Phasing was done wit SHAPEIT2^32^ and imputation was performed with Minimac3^33^. HRC reference release 1.1 was used as a reference panel. PLINK2 was used to run the GWAS, including SNPs with imputation score *>* 0.8 and MAF *<* 0.01, using logistic regression and the first five and the 7th MDS components as covariates. For the trio analysis, phasing and imputation were performed in the same way, GWAS was conducted without covariates.

Participants for the OCGAS study were recruited at five different participating sites of the National Institute for Mental Health at Johns Hopkins University School of Medicine, Brown Medical School, New York State Psychiatric Institute and College of Physicians and Surgeons at Columbia University, University of California Los Angeles (UCLA) School of Medicine, Massachusetts General Hospital and Harvard Medical School, National Institute of Mental Health, and Keck School of Medicine at the University of Southern California and was approved by each site’s IRB boards. Data was collected between 2007 and 2014 and is comprised of trios (affected proband and both parents) or a proband and an unaffected sibling. MD- or PhD-level clinical psychologists evaluated each individual using the Structured Clinical Interview for DSM-IV (SCID), the checklist of obsessions and compulsions from the Y-BOCS, refined to include the age of onset, age of offset, severity of each symptom, as well as the Y-BOCS scores for the worst episode (lifetime), further including course and treatment response variables. Children older than 8 years were assessed in the same way, but using the Kiddie-SADS instead of the SCID. Diagnostic status was assigned based on the consensus of two psychiatrists or psychologists independently evaluating each proband, and was further reviewed by one of five members of the JHU diagnostic consensus committee to ensure comparability across all five participating sites. The chance-corrected percent agreement between the diagnosticians for the diagnosis of OCD was *K* = 0.92; for age at onset of OCD, *K* = 0.88 (for age +/-5 years), and Pearson’s *r* = 0.71. Cases were required to meet DSM-IV criteria for OCD, with an onset of symptoms before the age of 18 (*mean* = 9.4 years; *SD* = 6.35). Individuals with a later age-of-onset, or with a diagnosis of schizophrenia, brain tumors, Huntington’s disease, Parkinson’s disease, Alzheimer’s disease, severe mental retardation, not permitting an evaluation of psychiatric disorders, or Tourette’s disorder or OCD occurring exclusively in the context of depression (secondary OCD) were excluded. Samples were genotyped on the Ilumina PsychChip array at USC.

### GWAS analysis 23andMe

#### Population

Samples of European ancestry were drawn from the research participant base of 23andMe, a private consumer genetics and research company. The cohort has been described in detail elsewhere ^34^. All participants provided informed consent, and answered online surveys. The cohort was selected from participant data available on 1st September 2017 and the analysis was reviewed and approved by a private institutional review board (www.eandireview.com). All individuals identified as cases have reported being diagnosed with OCD, while controls have reported not having been diagnosed with OCD. 30,167 OCD cases and 929,804 controls were included in the final GWAS. Of the cases, 10,808 (35.83%) were male and 19,359 (64.17%) were female. Of the controls, 443,336 (47,68%) were male and 486,468 (52,32%) were female. Of the cases, 7,040 (23.34%) individuals were below the age of 30, 11,087 (36.75%) between 30 and 45, 7,056 (23.39%) between 45 and 60, and 4,984 (16.52%) above the age of 60. Of the controls, 91,287 (9.82%) individuals were below the age of 30, 230,696 (24.81%) between 30 and 45, 257,793 between 45 and 60, and 350,028 (27.73%) above the age of 60.

#### Genotyping, QC, and imputation

Extraction of DNA and genotyping was performed by the National Genetics Institute (NGI), a CLIA licensed clinical laboratory and a subsidiary of Laboratory Corporation of America. Individuals were genotyped on four different genotype platforms. Two (V1, V2) platforms were variants of the Illumina HumanHap550+ BeadChip, including 25,000 custom SNPs, one platform (V3) was the Illumina OmniExpress+ BeadChip, with custom SNPs to increase overlap with V2, and one platform (V4) is in current use and a fully customized array. Individuals that failed to meet a 98.5% call rate were re-analyzed. Only individuals with *>* 97% European ancestry were included in the analysis. European ancestry was determined through an analysis of local ancestry using a support vector machine to classify individual haplotypes into one of 31 reference populations. Those classifications were then fed into a hidden Markov model (HMM) which accounts for incorrect assignments and switch errors, thereby giving probabilities for each reference population per window (100 SNPs). Simulated admixed individuals were then used to re-calibrate the HMM probabilities so the assigned ancestries were consistent with the simulated individuals. Publicly available datasets (Human Genome Diversity Project, HapMap, and 1000 Genomes) and 23andMe research participants with four grandparents from the same country served as the reference population. Identity-by-descent (IBD) estimation was used to define a maximal set of unrelated individuals for each analysis. Individuals who shared less than 700 cM IBD were defined as unrelated (e.g. approximately less related than first cousins).

The merged UK10K and 1000 Genomes Phase 3 panel was used for imputation. Finch, an internally developed tool, that implements the Beagle haplotype graph-based phasing algorithm, modified to separate the steps of graph construction and phasing, was used for phasing. Imputation was then performed as an estimated allele dosage averaged over a set of possible imputed haplotypes for each individual.

#### GWAS

Genetic association testing was performed using logistic regression, assuming an additive model for allelic effects, including age, sex, first five PCs, and the genotype platform as covariates. For imputed data, the imputed dosages rather than the best-guess genotypes were used. The association p-value was computed using a likelihood ratio test.

### GWAS meta-analysis

Before the above described datasets were combined in an OCD GWAS meta-analysis, each file was filtered and formatted in a way that they could be more easily combined. This involved aligning each SNP to the HRC genome ^35^ and excluding ambiguous SNPs and SNPs of likely poor quality. All variants had to meet the following criteria for inclusion: Case and control minor allele frequency *>* 1%, imputation-score *>* 0.8 and *<* 1.2. Strand-ambiguous A/T and C/G SNPs were removed if they had a frequency between 0.4 and 0.6. Was their frequency outside of that range, their frequency was compared to the frequency in the reference dataset. It was only retained if the frequency was either below 0.4 or above 0.6 in both, the OCD summary statistic and the reference dataset. If a variant did not overlap with the variants in the HRC reference, it was removed.

To combine the seven OCD case-control datasets we conducted an inverse variance weighted meta-analysis using METAL ^36^. The genome-wide significance threshold for the GWAS was set at a p-value of 5.0×10^*−*8^. For each individual GWAS as well as for the meta-analysis we calculated the genomic inflation factor (*λ*_1000_) to identify any residual population stratification of systematic technical artifact. We further calculated the linkage disequilibrium score (LDSC) regression intercept to evaluate the relative contribution of polygenic effects and residual artefacts such as population stratification and cryptic relatedness. For the analysis we used pre-computed LD scores from European-ancestry samples in the 1000 Genomes Projects (see web resources) and filtered our dataset on high-quality common SNPs with an INFO score *>*0.9. The influence of confounding factors was evaluated by comparing the estimated intercept of the LD score regression to one. One is the expected value of the LDSC intercept under the null hypothesis of no confounding (e.g. from population stratification). Heterogeneity in test-statistic across the seven datasets was assessed with Cochran’s *I*^2^ statistic. To identify any cohort in which the summary statistics significantly deviated from the rest of the cohorts, we performed sign test analyses for the top SNPs (inclusion threshold of *p* = 1.00×10^*−*4^, *p* = 1.00×10^*−*5^, *p* = 1.00x^*−*6^, and *p* = 5.00×10^*−*8^) between each cohort and leave-one out meta-analyses of the remaining 6 datasets.

### Gene-based tests

All gene-based tests were conducted using MAGMA v1.08^37^. Gene units were defined using three separate models: 1) the standard MAGMA model based on proximity, with a SNP falling within the general gene region, here defined as 35kb upstream of the transcription start site to 10kb downstream of the transcription start site; 2) a model where genes units are formed across 13 different brain tissues from GTEx by defining SNPs as eQTLs for genes in each tissue in question ^38^; 3) a model defined by SNPs linked to gene regions via 3D chromatin interactions across eight brain tissue datasets ^39^. All MAGMA gene-based tests used the following inclusion filters: MAF *≥* 0.01, INFO *≥* 0.9, and additional MAGMA parameters ‘use=SNP,P ncol=Neff_half’. The ‘use=SNP,P’ means that the SNP IDs are derived from the ‘SNP’ column of input daner files, and that p-values were derived from the ‘P’ column of these files. The ‘ncol=Neff_half’ refers to the definition of N per SNP as the effective sample size and can be defined as 4*Nca*Nco/(2*(Nca+Nco)) in a single cohort. A Bonferroni threshold for significance was set based on the total number of tests performed across these three models combined (45,664) and was equal to 0.05 / 45,664 = 1.09×10^*−*6^.

### Tissue / celltype enrichment analyses

We followed a tissue / celltype enrichment analysis protocol recently described in Bryois et al. ^40^. Consistent with this we utilized the codebase for this analysis, and in particular the sets of genes that mark different highlighted tissue and celltype datasets, that were made available by the manuscript authors at https://github.com/jbryois/scRNA_disease.

We selected 3 datasets that had been preprocessed by Bryois et al. ^40^ for inclusion in this analysis. The first features tissue-specific gene expression data derived from GTEx ^41^, with a total of 37 tissues represented in the data. The second and third are derived from Zeisel et al. ^42^, and represent broad celltype groups across the entirety of the mouse nervous system, along with a high resolution single celltype map of the same data. In Zeisel et al. ^42^, a total of 39 broad celltype groups and 265 individual celltypes are represented.

We followed the analysis protocol described in Bryois et al. ^40^ for the analyses of 37 tissues from GTEx and 39 broad celltype groups from Zeisel et al. ^42^, and utilized a simplified approach for the analysis of each of the 265 individual celltypes from Zeisel et al. ^42^. For the tissue and broad celltype group datasets, we conducted the full protocol from Bryois et al. ^40^ which included analyzing tissue/celltype enrichment using both LDSC ^43^ and MAGMA ^44^. We only considered a tissue / celltype significantly enriched if the FDR-adjusted p-value was less than 0.05 in both the LDSC and MAGMA-based tests. Due to the high computational demands of testing 265 individual celltypes across the mouse nervous system with LDSC, we limited the protocol to only include the MAGMA assessment. We considered a celltype significant if it had an FDR-adjusted p-value less than 0.05.

All statistical analysis downstream of LDSC and MAGMA (namely, p-value adjustment) was done using R v4.0.0, and all plotting was done inside of R v4.0.0 using the package ggplot2 v3.3.2. FDR p-value adjustments were performed using the R function p.adjust(method=“fdr”). Bonferroni p-value adjustments were performed using the R function p.adjust(method=“Bonferroni”).

### Multivariable GWAS with MTAG

We used MTAG ^45^ to conduct multivariable GWAS analyses for the OCD samples. MTAG combines related traits into a meta-analysis by leveraging the shared heritability among the different traits. For this purpose we included the same summary statistics that also formed the basis for the meta-analysis using METAL, i.e., the freeze 1 data from the original meta-analysis together with the NORDiC samples (freeze1+NORDiC, see above), the iPSYCH dataset, and the 23andMe dataset. But unlike with the METAL-based meta-analysis, the MTAG analysis results in three subgroup-specific estimates (i.e., freeze1+NORDiC, iPSYCH, and 23andMe) that gain power by exploiting the high shared heritability across all subgroups compared to separate subgroup specific analysis. Through this approach we aimed to address potential concerns about heterogeneity in our phenotyping strategies for the three individual sub datasets (see below in the discussion). We performed maxFDR analyses to approximate the upper bound on the FDR of MTAG results.

### SNP-based heritability and genetic correlation with other disorders and traits

LDSC ^43^ was used to evaluate the contribution of all included SNPs on the variance in liability to OCD (SNP-based heritability). The analysis was performed using pre-computed LDscores from samples restricted to European-ancestry in the 1000 Genomes Project ^43;46^, filtered for SNPs included in the HapMap3 reference panel ^47^. SNP-heritability was estimated based on the slope of the LD score regression, with heritability on the liability scale calculated assuming a 3% population prevalence of OCD. SNP-heritability was calculated for the whole OCD sample as well as for a) clinical samples (NORDiC-SWE, NORDiC-NOR, and freeze1), b) samples not primarily ascertained for OCD (iPSYCH), and c) self-reported biobank samples (23andMe). Cross-trait LDSC regression was used to estimate the genetic correlations of OCD with 74 other phenotypes (see Supplementary Table S7 for an overview of all tested traits). LDSC estimates the genetic correlation between two traits by regressing the product of the Z-scores from GWASs on the LD score which represents the genetic co-variation between the two traits based on all polygenic effects captured by the included SNPs. Again, European-ancestry LD-scores from the 1000Genomes project, restricted to SNPs included in the HapMap3 reference panel, were used. Cross-trait genetic correlations were calculated for the whole OCD sample, for the same three sub-samples used in the SNP heritability analysis, and for the three MTAG analyses.

## Results

### GWAS meta-analysis

We conducted a GWAS meta-analysis of seven European-ancestry case-control cohorts, including a total of 37,015 OCD cases and 948,616 controls with 6,829,695 autosomal SNPs. Five datasets have been published in previous OCD-GWASs - the OCGAS and IOCDF-GC cohorts have been included in the first OCD GWASs ^16;18;19^, while the NORDiC-SWE, NORDiC-NOR, and iPSYCH cohorts have been included in a recent preprint ^20^. For this GWAS, additional samples from 23andMe (30,167 cases and 929,804 controls) were added to these previously published datasets. The QQ-plot (Figure 1B) and Lambda (*λ* = 1.241; *λ*_1000_ = 1.003) of the meta-analysis was in accordance with the assumption of polygenicity and did not show any evidence for residual stratification effects. 15 independent SNPs exceeded the genome-wide threshold for significance (See Figure 1, Supplementary Figures S2-S16 for regional association plots and forest-plots). The genome-wide significant SNPs were located on chromosomes 1, 2, 3, 4, 5, 6, 8, 11, 12, 15, and 21. The SNP with the lowest p-value was rs10877425 on chromosome 12 (P = 1.14×10^10^) and there were an additional 27 SNPs that were genome-wide significant in this locus.

**Fig. 1.**
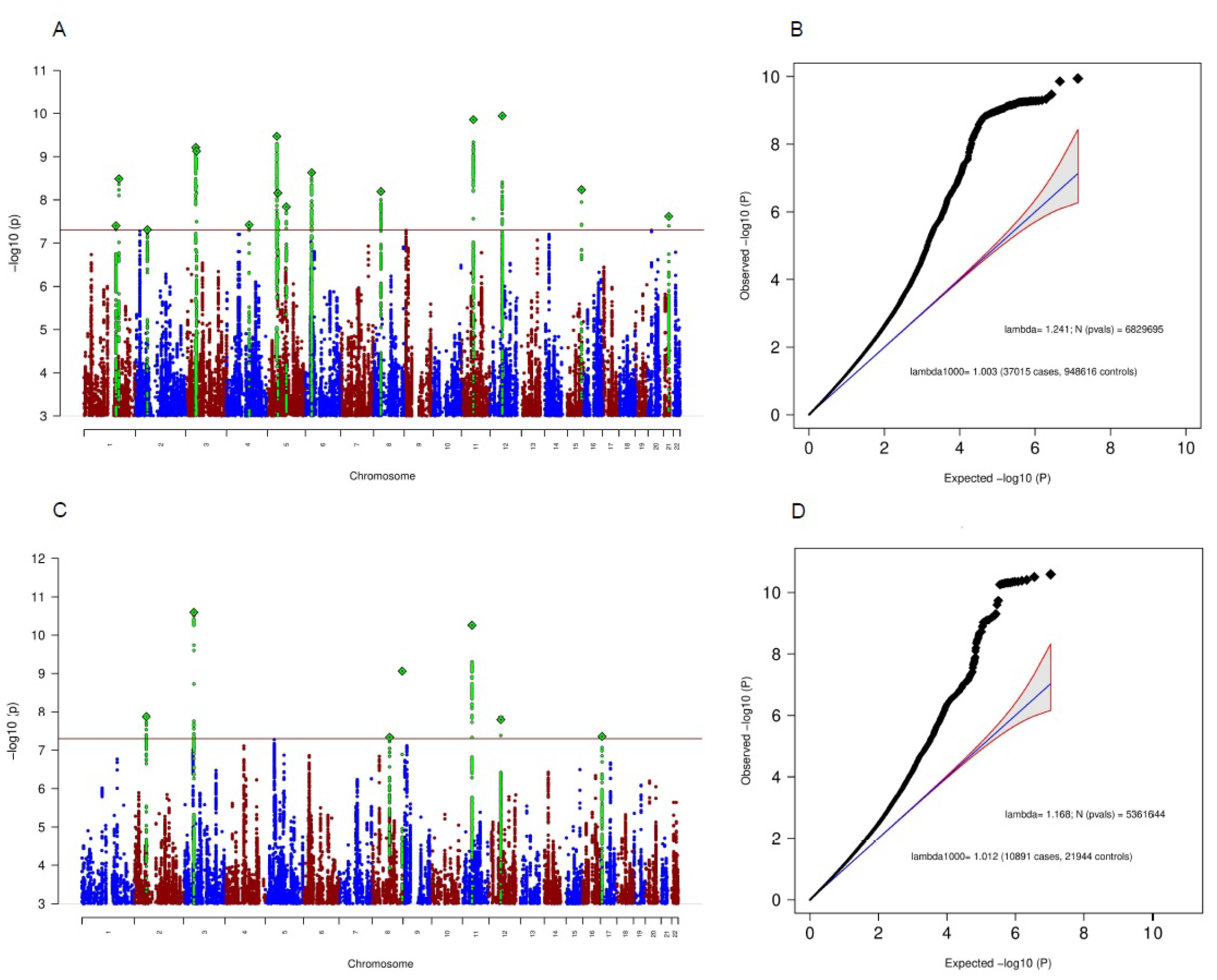
Genome-wide association results for the OCD meta-analysis (37.015 OCD cases and 948.616 controls; Manhattan plot in (A) and quantile-quantile plot in (B)) and for the MTAG analysis with freeze1+NORDiC as the primary phenotype (Manhattan plot in (C) and quantile-quantile plot in (D)). The x-axis in (A) and (C) shows the position in the genome (chromosomes 1-22), while the y-axis represents –log10(p) values for the association of variants with OCD, The horizontal red line represents the threshold for genome-wide significance. Each dot represents one SNP that was tested in the GWAS, with green diamonds indicating the lead SNP in regions harboring a genome-wide significant SNP. In (B) and (D) the expected -log10(p) under the null is plotted against the observed -log10(p), qq-plots (B) and (D) belong to Manhattan lots (A) and (C), respectively. The shading indicates 95% confidence region under the null. Lambda indicates the genomic inflation factor.

### Gene-based tests

Gene-based tests were conducted using MAGMA v1.08 to determine if, based on various criteria, any single protein-coding genes carried a load of common variation in OCD relative to controls that passed the Bonferroni threshold for significance. We conducted three sets of tests, each one linking common variation to individual genes via different means. The first was the standard MAGMA approach, considering SNPs as part of the signal for a given gene if it falls within the region 35kb upstream of the gene transcription start site to 10kb downstream of the transcript stop site (18,048 tests total). The second, e-MAGMA, considered SNPs as members of a gene if they were called as eQTLs in at least one brain tissue type assessed in GTEx (9240 tests total). The third, h-MAGMA, considered SNPs as members of a gene if they fell within gene exons or promoters, or alternatively if they were in distal intronic/intergenic regions but overlapped with a chromatin interaction detected in fetal or adult brain that linked back to the gene body (18,376 tests total). The threshold for significance was set based on the sum of the described tests performed (0.05/45,664 = 1.1×10^*−*6^).

We identified a total of 101 tests that passed the Bonferroni threshold for significance, mapping back to a total of 79 distinct protein-coding genes (Figure 2). Only one of these genes, *WDR*6, was observed as significant across all tests. Results from c-MAGMA, e-MAGMA and h-MAGMA captured overlapping and unique significant single-gene results (Figure 3). In some instances, the extended versions of MAGMA (e-MAGMA and h-MAGMA) provide additional context to the gene-based signal. For example, h-MAGMA suggests that the significantly associated gene *HCN* 1 is concentrated within promoter, exonic and chromatin-interacting loci. In another example, only e-MAGMA implicates a set of 6 genes within a region on chromosome 6, suggesting that the eQTLs that are a part of this signal could not be captured via simple gene overlap or 3D chromatin interactions. All gene-based test results are provided in Supplementary Table S2-S5.

**Fig. 2.**
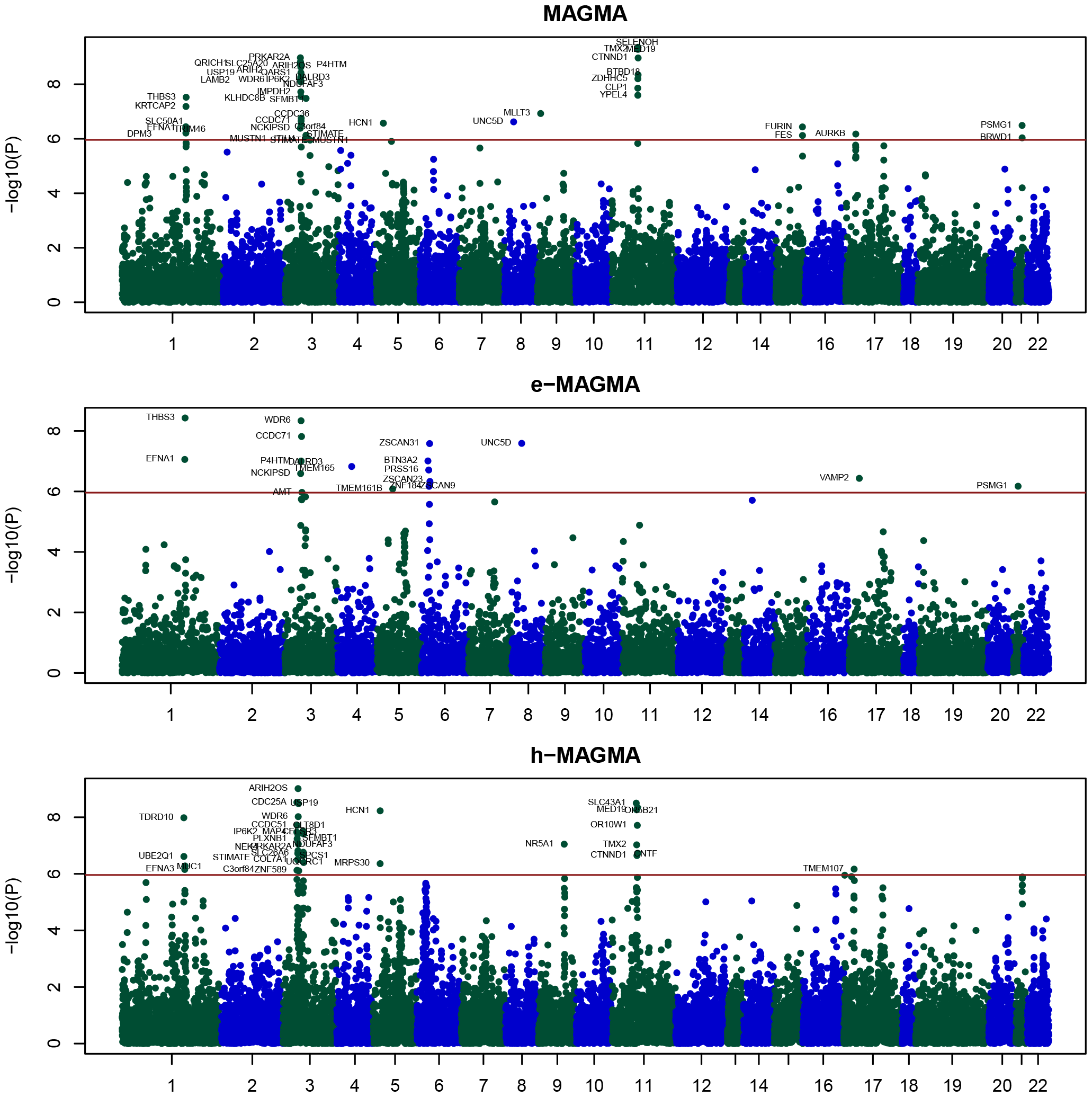
Manhattan plots for single-gene results from MAGMA, e-MAGMA and h-MAGMA analyses. Single gene result significance levels (−log10 p-values) are shown for MAGMA (top), e-MAGMA (middle) and h-MAGMA (bottom) approaches. A total of 79 distinct single protein coding genes passed the significance threshold shown in at least 1 of 3 sets of tests.

**Fig. 3.**
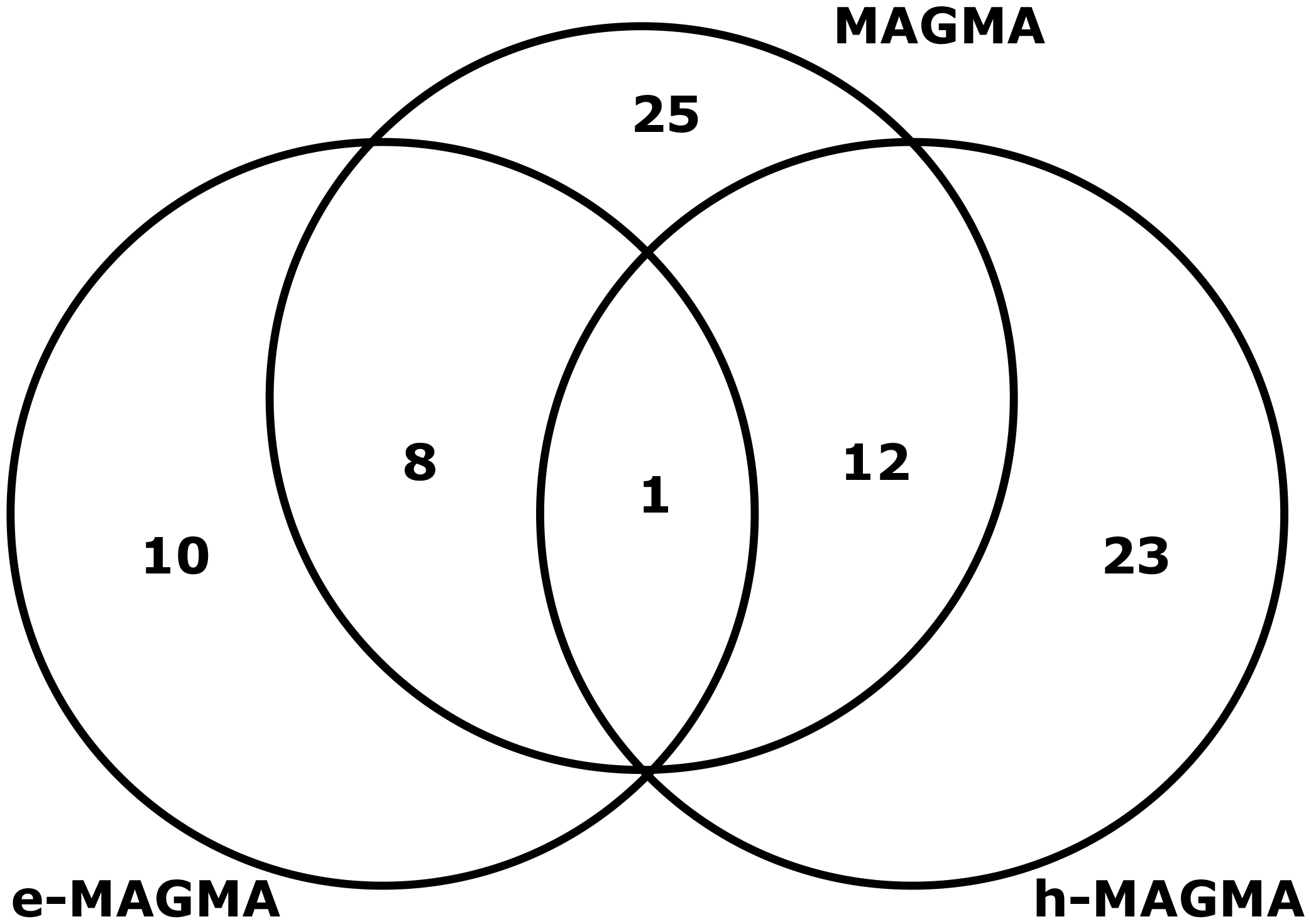
Overlaps of genes called as significant in MAGMA, e-MAGMA and h-MAGMA. Overlaps between the three methods for conducting gene-based tests using GWAS summary statistics are depicted via a venn diagram, with the numbers representing the number of genes called as significant across methods.

### Tissue / celltype enrichment analyses

In our enrichment analyses focused on broad tissues as defined by GTEx ^41^ we identified five different tissues that carried a significant amount of OCD heritability enrichment. All tissues were found within the brain, and included the Anterior cingulate Cortex, Frontal Cortex, Amygdala, and Hypothalamus (Figure 4).

**Fig. 4.**
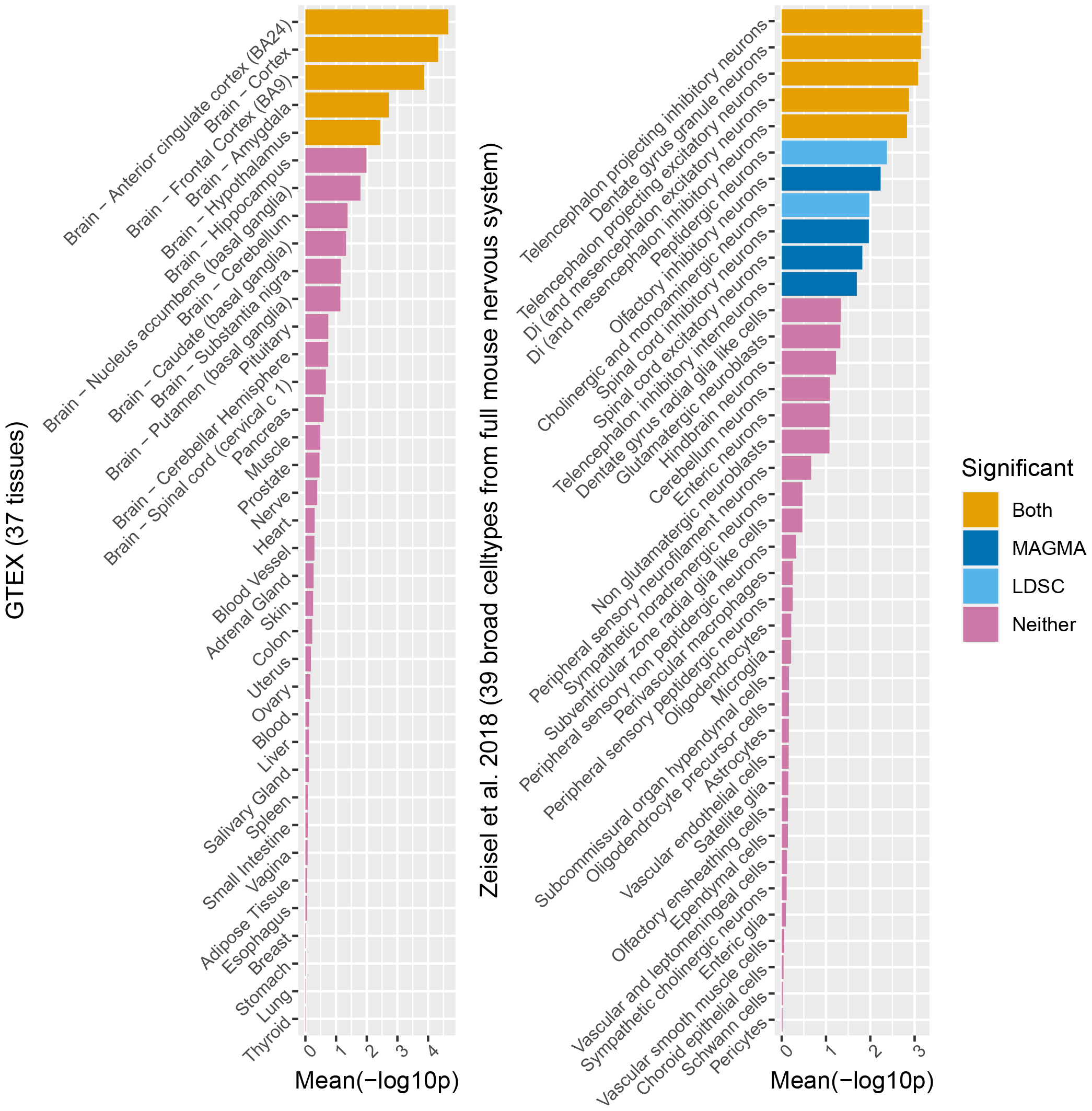
OCD heritability enrichment p-values across tissue/celltype expression profiles. Test result p-values are shown for heritability enrichment tests with genes representing expression profiles of 37 different tissues from GTEx (left) and 39 broad nervous system celltypes from mice (right) The p-value shown for each result is averaged between MAGMA and LDSC approaches, and is considered to be significant only if FDR-adjusted p < 0.05 in both. A total of 5 broad tissue types from GTEx (all from brain), and a total of 5 broad neuronal celltypes from Zeisel et al. are significantly enriched for OCD heritability based on these criteria.

Enrichment analyses focused on broad celltypes were carried out in the same manner as tissue enrichment analyses and identified a total of five broad celltypes that are significant based on the methods used (see methods). Significant results included telencephalon projecting excitatory and inhibitory neurons, di- and mesensephalon exicitatory and inhibitory neurons, and dentate gyrus granule neurons (Figure 4).

We conducted these enrichment analyses on a total of 265 fine-grain celltypes from the mouse nervous system ^42^, and identified a total of 44 that survived FDR correction (Supplementary Table S10). We formed 35 groups of single cell types where N *≥* 2 based on the ‘Description’ column from the table in http://mousebrain.org/celltypes/. We then took the mean -log10 p-value from each group, and performed FDR correction on these aggregated statistics in order to determine if any of these groups carried a significant excess. We identified a total of seven significant classifications, with key results including D1 and D2 medium spiny neurons, along with excitatory neurons from hippocampus C1, and cerebral cortex (Figure 5).

**Fig. 5.**
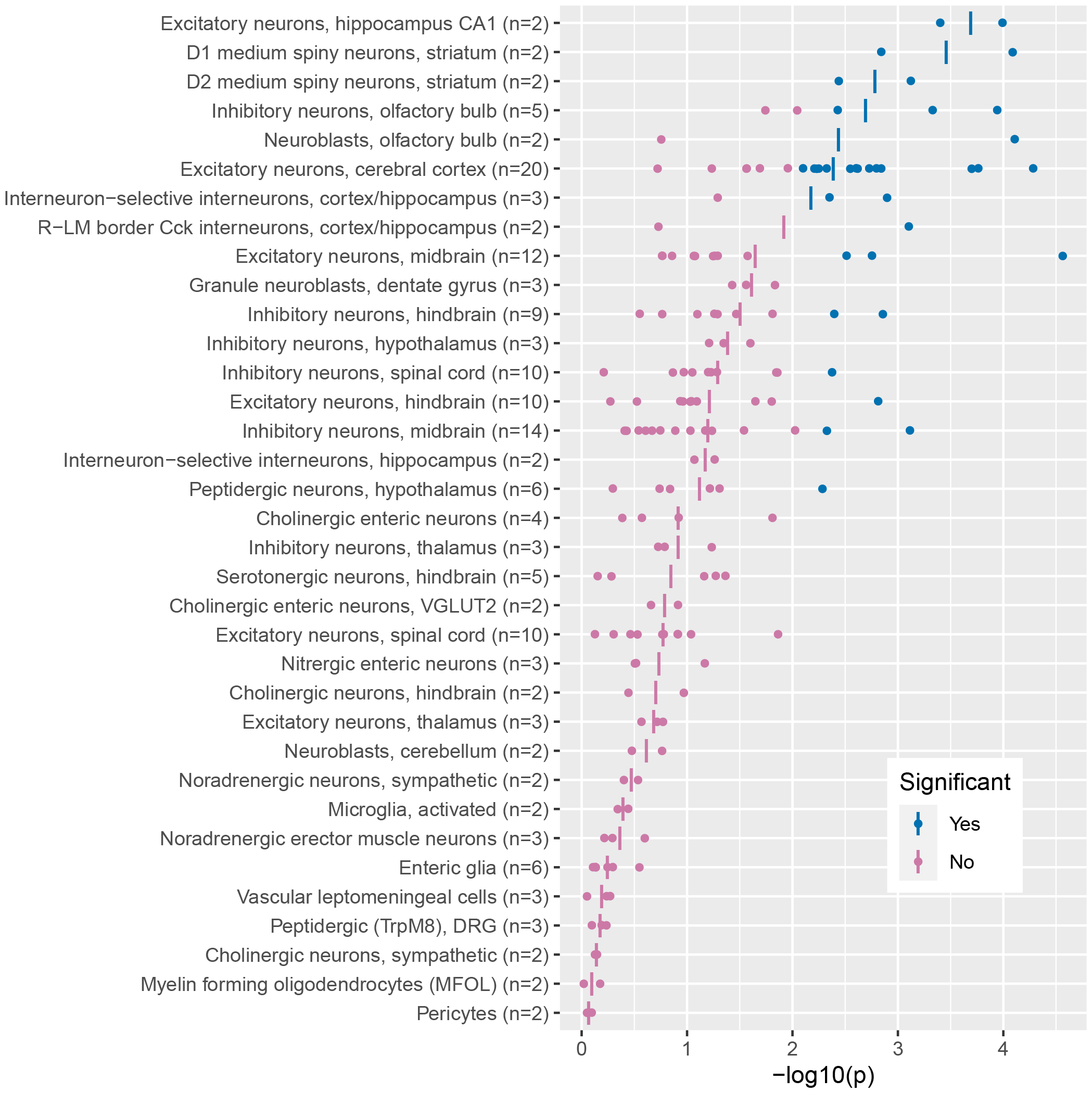
Heritability enrichment p-values between OCD and neuronal celltype groups. P-values for OCD heritability enrichment across 265 celltypes from Zeisel et al. were generated, and grouped into 35 groups of single cell types where n>=2 based on the ‘Description’ column from the table in http://mousebrain.org/celltypes/. We first tested each of the 265 celltypes and found that a total of 44 were significant at an FDR < 0.05. We then took the mean -log10 p-value from each of the 35 groups and found that of these a total of 7 were significant at FDR < 0.05. We indicate in blue the single celltypes and overall celltype groups that are significant after FDR adjustment.

### MTAG analyses

Leveraging the shared heritability among the three characteristically differing OCD sub-samples (freeze1+Nordic, iPSYCH, 23andMe), we performed three separate multi-trait analysis with MTAG, with the aim to extract specific genetic underpinnings characteristic of a) clinically ascertained OCD cases (freeze1+Nordic) b) OCD cases comorbid with another psychiatric disorder (iPSYCH), and c) self-report OCD status (23andMe).

For freeze1+NORDiC (clinically ascertained) we identified seven genome-wide significant associated SNPs (see figure 1C for the Manhattan plot and figure 1D for the qq-plot). Four of these SNPs overlap with the original GWAS meta-analysis of OCD, one genetic region overlaps with the MTAG analysis focusing on the iPSYCH sample (comorbid ascertainment) and the MTAG analysis focusing on the 23andMe sample (self-report), one SNP overlaps with the MTAG analysis focusing on the iPSYCH sample, and one is unique for clinically ascertained OCD cases. Max FDR was calculated at 0.0371. For iPSYCH we identified 15 significant SNPs, of which 11 regions (the most significant SNP in a region varied) overlap with the original GWAS meta-analysis, one overlaps with the other two MTAG analyses, and four SNPs overlap with one of the other two MTAG analyses. Max FDR was calculated at 0.0154. For 23andMe we identified 18 genome-wide significant SNPs, of which 12 regions overlapped with the original GWAS meta-analysis, three overlapped with at least one of the other MTAG analyses and three were uniquely significant in this analysis. Max FDR was calculated at 0.0035.

A total of five loci were significant in all three runs of MTAG (one locus was tagged by two different SNPs: rs13262595 in freeze1+NORDiC and rs4129585 in iPSYCH and 23andMe) and four of these were significant in the original meta-analysis of these samples. Overall p-values of the lead SNPs appeared more significant in the MTAG analyses when compared to the original GWAS meta-analysis. See Supplementary Table S6 for an overview of all significant SNPs in the MTAG analyses compared to the original GWAS meta-analysis.

### SNP-based heritability and genetic correlation with other disorders and traits

The SNP-based heritability of OCD, calculated using LD-score regression (LDSC), was estimated to be 0.085 (*SE* = 0.004) for the whole sample, assuming a 3% population prevalence. Subdividing the sample into the three subsets (freeze1+NORDiC, iPSYCH, 23andMe) and keeping the population prevalence at 3% yielded liability scale heritability estimates of 0.321 (*SE* = 0.039), 0.25 (*S*E = 0.045), and 0.081 (*SE* = 0.005), respectively. Genetic correlations between the three sub-samples were high, reaching *rg* = 0.88 (*SE* = 0.13, *P* = 2.01×10^*−*12^) between freeze1+NORDiC and iPSYCH, *rg* = 0.64 (*SE* = 0.06, *P* = 5.56×10^*−*29^) between freeze1+NORDiC and 23andMe, and *rg* = 0.85 (*S*E = 0.09, *P* = 1.52×10^*−*21^) between iPSYCH and 23andMe.

Of the genetic correlations between OCD and 74 psychiatric, personality, psychological, substance-use, neurological, cognition, socioeconomic status, autoimmune, cardiovascular, anthropomorphic, and fertility traits, 40 exceeded the FDR-corrected significance threshold (Figure 6, Supplementary Table S7). OCD positively correlated with all psychiatric disorders, with especially high correlations with ANX, depressive disorder/MDD, PTSD, AN, and TS. Also, alcohol- and nicotine dependence correlated positively with OCD, as well as several other smoking related phenotypes. While intelligence and educational attainment correlated negatively, OCD correlated positively with memory. Moreover, OCD was significantly positively genetically correlated with neuroticism (especially with the worry sub-cluster), conscientiousness, loneliness, and tiredness, while the genetic correlation with household income, subjective well-being, self-rated health, and sleep duration was negative. Notable are also the significant negative correlations with the three autoimmune disorders ulcerative colitis, Crohn’s disease, and inflammatory bowel disease and a positive correlation with adult-onset asthma. Moreover, OCD showed a positive genetic correlation with ALS and childhood maltreatment and a negative correlation with BMI and age at first birth. Correlations with other neurological, cardiovascular, anthropomorphic and fertility traits did not demonstrate a significant correlation with OCD.

**Fig. 6.**
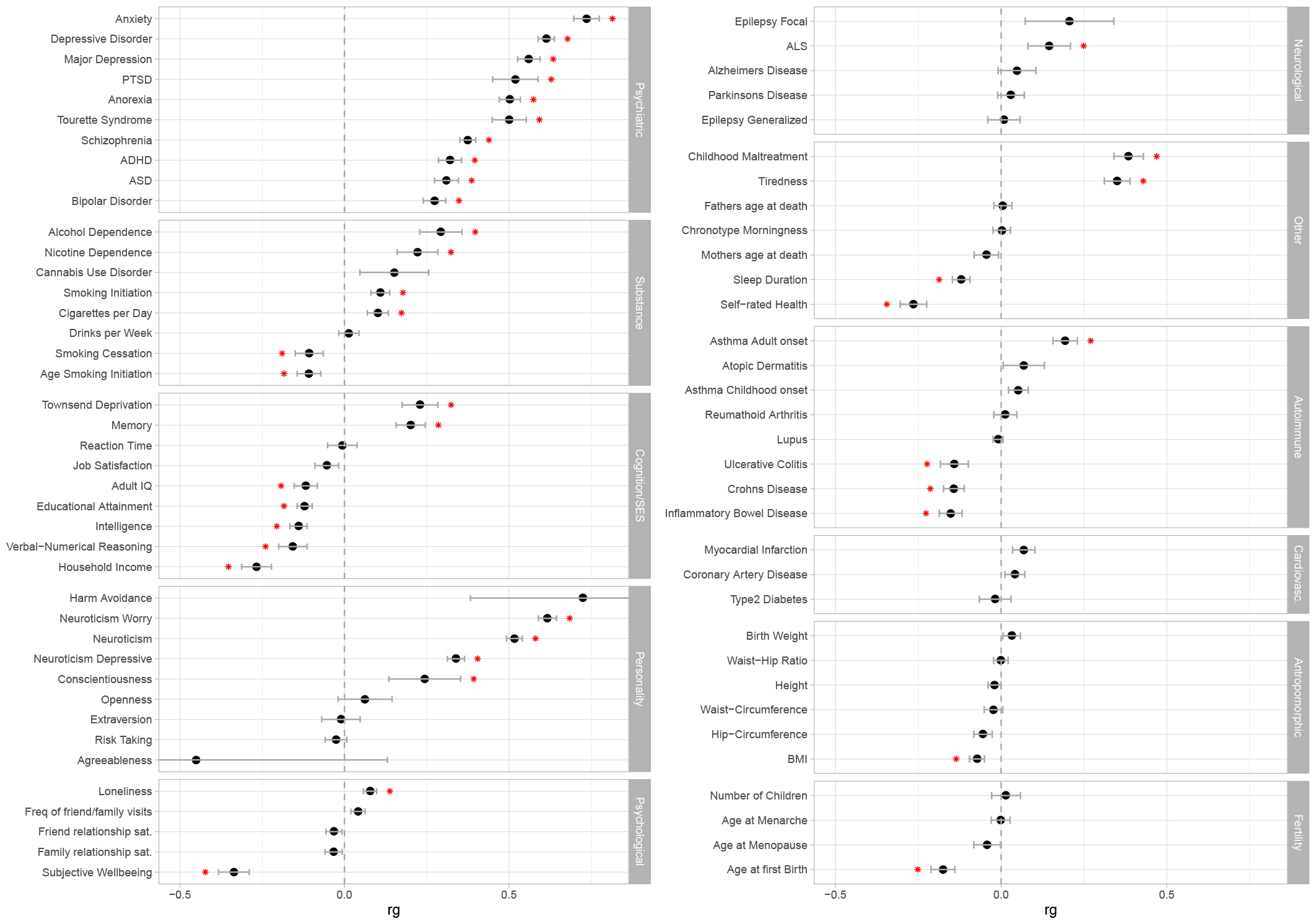
Genetic correlations (rg) between OCD and 74 behavioral, cognitive, psychiatric, neurologic, immunologic, metabolic, and anthropomorphic phenotypes. Error bars represent standard errors and red asterisks indicate significant associations after FDR correction for multiple testing.

We further examined the genetic correlations of the three OCD sub-samples, freeze1+NORDiC, iPSYCH, and 23andMe, and the 74 traits (Supplementary Figure S17, and Supplementary Table S8). All three sub-samples showed significant genetic correlations in the same direction for most psychiatric disorders (except ADHD and PTSD), neuroticism, neuroticism worry sub-cluster, and subjective well-being. Across all trait-categories, the correlations of the clinically ascertained sub-sample were generally lower than the correlations of the other two sub-samples, with the exception of the three autoimmune disorders ulterative colitis, Crohn’s disease, and inflammatory bowel disease, as well as several anthropomorphic traits. Also for the substance-use phenotypes we found divergence in the correlation pattern - only the self-reported samples show a negative correlation with IQ, educational attainment, verbal-numerical reasoning and household income.

We also repeated the genetic correlation analyses for the OCD summary statistics resulting from the three MTAG analyses (Supplementary Figure S18, and Supplementary Table S9). As expected, correlations with all three OCD MTAG summary statistics were more uniform than the correlations with the individual sub-samples (Supplementary Figure SS17) and similar to the genetic correlation estimates of the over all OCD GWAS meta-analysis (Figure 6).

## Discussion

Here we report results from the largest OCD GWAS to date. We find support for 15 independent genome-wide significant loci (14 new) and 79 protein coding genes in genebased tests. Tissue enrichment analyses implicate multiple cortical regions, the amygdala and hypothalamus while cell type analyses yielded 12 cell types linked to OCD (all neurons). The SNP heritability of clinically ascertained samples (freeze1+NORDiC; 0.318) was comparable to samples ascertained for a different co-morbid disorder (iPSYCH; 0.254) but higher than self-reported OCD samples (23andMe; 0.081), although all three sub-samples were highly correlated. OCD was genetically correlated with 40 disorders or traits – positively with all psychiatric disorders and negatively with BMI, age at first birth and multiple related autoimmune diseases. Overall, OCD appears to be on a trajectory similar to other psychiatric disorders in terms of common-variant discovery as sample size grows.

We replicated the one previously described ^20^ genome-wide significant locus for OCD at (rs2581789 at 3p21.1) and refined this hit (rs2564930 is now the lead SNP; see Supplementary Figure S6). As described before ^20^, this region has previously been associated with a broad range of other psychiatric disorders and related traits, including schizophrenia ^48^, well-being ^49^, and the worry-subcluster of neuroticism ^50^. This variant was genome-wide significant in all three MTAG analyses. We also describe, for the first time, an association between OCD and the major histocompatibility complex (MHC) region of chromosome 6 (Supplementary Figure S7). The lead SNP, rs9265969, is just 6kb away from the HLA-B gene. This is notable for multiple reasons. First, much attention has been devoted to the possible alterations of the immune system in OCD ^51^ and there is a familial link between autoimmune diseases and OCD ^52^. Second, other psychiatric disorders (e.g., schizophrenia and bipolar disorder) have shown association with the MHC ^48;53^, with schizophrenia’s association with the MHC locus arising in substantial part from many structurally diverse alleles of the complement component 4 (C4) genes ^54^. It will be interesting to see whether imputation of C4 alleles using SNP data uncovers an association with OCD, or like bipolar disorder, reveals no association with C4.

We further found OCD to be associated with rs4702 in the 5’ UTR of the gene *FURIN* (Supplementary Figure S9). This variant is a known eQTL, with the OCD risk variant (G) associated with decreased expression in the brain ^55^. Of note, the same variant has previously been associated with schizophrenia and bipolar disorder with the same direction of effect ^48;53^. There were 12 other genome-wide significant hits, including 2 intergenic loci, 3 that are multigenic, and 7 where a single gene is contained within the genome-wide significant region (these 7 genes are *HCN1, LINC00970, UNC5D, LINC02106, BRWD1, SLC39A8* and *LINC01122*). Little is known about the 3 *LINC* RNA genes, although *LINC01122* is highly expressed in the brain relative to other tissues in GTEx. *HCN1* encodes a hyperpolarization-activated cation channel that contributes to spontaneous rhythmic activity in both heart and brain and rare variants in this gene have been associated with epilepsy ^56^. *UNC5D* is a receptor for netrin-1 and is thought to play a role in axon guidance by mediating axon repulsion of neuronal growth cones in the developing nervous system ^57^. *BRWD1* is thought to act as a transcriptional regulator involved in chromatin remodeling and is located within the Down syndrome region-2 on chromosome 21^58^. *SLC39A8* encodes a plasma membrane transporter mediating the cellular uptake of zinc and manganese, two divalent metal cations important for development, tissue homeostasis and immunity ^59^. Of note, in a small study zinc was used as an adjuvant to fluoxetine and produced improved response ^60^.

We performed three sets of gene-based tests using MAGMA (standard MAGMA ^44^, e-MAGMA ^61^ and h-MAGMA ^39^) to link common variation to protein-coding genes. A total of 79 genes were implicated, including 21 genes that were found with at least two versions of MAGMA, and one gene (*WDR6*) that was found with all three methods (see Figure 2 and Figure 3). *WDR6* is a member of the WD repeat protein family which is implicated in cell growth arrest ^62^. We further performed three sets of multivariable GWAS (MTAG) to obtain ascertainment specific estimates while gaining power through leveraging the shared heritability among the subgroups. A total of five loci were significant in all three runs of MTAG (see Supplementary Table S6) and four of these were significant in the standard GWAS. The one MTAG-significant locus that was not significant in the standard GWAS contains *TSNARE*1, which encodes a protein thought to play a crucial role in intracellular protein transport and synaptic transmission ^63^ and has been repeatedly associated with schizophrenia and other neuropsychiatric traits ^64^.

We used MTAG ^45^ to conduct multivariable GWAS analyses for the OCD samples. MTAG combines related traits into a meta-analysis by leveraging the shared heritability among the different traits. For this purpose we included the same summary statistics that also formed the basis for the meta-analysis using METAL, i.e., the freeze 1 data from the original meta-analysis together with the NORDiC samples (freeze1+NORDiC, see above), the iPSYCH dataset, and the 23andMe dataset. But unlike with the METAL-based meta-analysis, the MTAG analysis results in three subgroup-specific estimates (i.e., freeze1+NORDiC, iPSYCH, and 23andMe) that gain power by exploiting the high shared heritability across all subgroups compared to separate subgroup specific analysis. Through this approach we aimed to address potential concerns about heterogeneity in our phenotyping strategies for the three individual sub datasets (see below in the discussion). We performed maxFDR analyses to approximate the upper bound on the FDR of MTAG results.

Our tissue enrichment results are consistent with other psychiatric disorders in that only brain tissues were found to be enriched for OCD heritability (Figure 4). Specific tissues were the anterior cingulate cortex, frontal cortex, amygdala, and hypothalamus, although a number of other brain regions approached significance, including sub-components of the striatum. Imaging studies as well as neuropsychological and treatment studies have implicated a cortico–thalamo–striatocortical (CTSC) circuit in the pathophysiology of OCD ^14^. Therefore, while there is some overlap in brain regions implicated by OCD genetics and imaging, larger studies and more precise brain tissue expression data are needed to clarify this relationship. We next sought to identify cell-types enriched for OCD heritability using single-cell RNA sequencing data. As shown in Figure 5, a total of 12 cell types reached significance, including multiple types of cortical neurons and both dopamine D1 and D2 receptor-expressing medium spiny neurons (MSNs) of the striatum. MSNs are a special type of GABAergic inhibitory cells representing 95% of neurons within the human striatum, a central structure of the CTSC circuit mentioned above.

With regards to the difference in reported liability-scale heritability for the three sub samples there could be multiple reasons. For example, on average our freeze1+NORDiC sub sample (as well as the iPSYCH sub sample) likely includes more severely and more chronically affected individuals in contrast to the 23andMe sub sample (which might represent on average healthier individuals within the OCD spectrum). The effect of such a difference on heritability estimates and variance explained has been reported before ^65;66^.

Our LDSC results revealed that OCD was genetically correlated with 40 disorders or traits (Figure 6). This included positive correlations with all psychiatric disorders, most notably anxiety, depression, PTSD, anorexia and Tourette syndrome. These results are consistent with prior studies of OCD as well as known comorbidities. Negative correlations were found for BMI and age at first birth (as described before ^20^) as well three related autoimmune disorders (ulcerative colitis, Crohn’s disease and inflammatory bowel disease), which is perhaps surprising given the association with the MHC and the familial link between autoimmune diseases and OCD.

In summary, we report 15 independent genome-wide significant loci associated with OCD. The GWAS meta-analysis implicates the MHC region, *FURIN* and other biologically informative genes as important contributors to the etiology of OCD. The results also highlight strong overlap with the genetics of OCD-related disorders and traits in the population, encouraging a multi-faceted view on the genetic underpinnings of OCD and potentially a continuum of genetically distinctive sub-entities. Overall, we have begun laying the groundwork through which the biology of OCD and related phenotypes will be described.

## Data Availability

GWAS summary statistics will be made available on the iPSYCH website (https://ipsych.dk/en/research/downloads), excluding 23andMe data.
The full GWAS summary statistics for the 23andMe discovery data set will be made available through 23andMe to qualified researchers under an agreement with 23andMe that protects the privacy of the 23andMe participants. Please visit https://research.23andme.com/collaborate/#dataset-access/ for more information and to apply to access the data.

## FUNDING ACKNOWLEDGEMENTS

The iPSYCH team was supported by grants from the Lundbeck Foundation (R102-A9118, R155-2014-1724, and R248-2017-2003), NIH/NIMH (1R01MH124851-01 to A.D.B.) and the Universities and University Hospitals of Aarhus and Copenhagen. The Danish National Biobank resource was supported by the Novo Nordisk Foundation. High-performance computer capacity for handling and statistical analysis of iPSYCH data on the GenomeDK HPC facility was provided by the Center for Genomics and Personalized Medicine and the Centre for Integrative Sequencing, iSEQ, Aarhus University, Denmark (grant to A.D.B.). A.D.B. was also supported by the EU’s HORIZON-HLTH-2021-STAYHLTH-01programme, project number 101057385: Risk and Resilience in Developmental Diversity and Mental Health (R2D2-MH). The NORDiC study was supported by NIH grant R01MH110427 (PI: Crowley), NIMH R01 MH105500 (PI Crowley); the Swedish Research Council (grant numbers 2015-02271, 2018-02487) (PIs: Mataix-Cols and Rück), CIMED and Region Stockholm (PI: Rück). The computation was performed on resources provided by SNIC through Uppsala Multidisciplinary Center for Advanced Computational Science (UPPMAX) under Project sens2018605.

We would like to thank the research participants and employees of 23andMe for making this work possible. Participants provided informed consent and participated in the research online, under a protocol approved by the external AAHRPP-accredited IRB, Ethical Independent Review Services (EI Review). Participants were included in the analysis on the basis of consent status as checked at the time data analyses were initiated.

## DISCLOSURES

David A. Hinds and Chao Tian are employed by and hold stock or stock options in 23andMe, Inc. ADB has received speaker fee from Lundbeck. JH has received lecture honoraria as part of continuing medical education programs sponsored by Shire, Takeda and Medice. DMC receives royalties for contributing articles to UpToDate, Wolters Kluwer Health, and personal fees for editorial work from Elsevier, all unrelated to the current work.

All other authors report to conlficts of interests.

## DATA AVAILABILITY

GWAS summary statistics will be made available on the iPSYCH website (https://ipsych.dk/en/research/downloads), excluding 23andMe data. The full GWAS summary statistics for the 23andMe discovery data set will be made available through 23andMe to qualified researchers under an agreement with 23andMe that protects the privacy of the 23andMe participants. Please visit https://research.23andme.com/collaborate/dataset-access/ for more information and to apply to access the data.

**Table 1.** Top SNP results of OCD meta-analysis showing all significant SNPs, and corresponding chromosome (CHR), basepair position (BP), effect allele and non-effect allele (A1/A2), frequency of allele A1 for cases (FRQ cases), frequency of allele A1 for controls (FRQ controls), log odds ratio (log(OR) Meta), and p-value for association results (P Meta). Also listing protein coding genes and/or microRNAs in a LD region of *r*^2^=0.6 around the lead SNP (±50 kb) in brackets distance to index SNP in kb (Genes). If more than one gene is contained within that region, only selected genes with a known association with OCD or a related disorder together with the number of genes (in brackets) within that region are listed in the table, while the remaining genes are listed at the end of this description. Also listing FRQ cases, log(OR), and P separately for freeze1+NORDiC, including 4417 OCD cases from NORDiC-NOR, NORDiC-SWE, and freeze1 (OCGAS, IOCDF-GC, and IOCDF-GC_trios); for iPSYCH (including 2678 OCD cases), and for 23andMe (including 30167 OCD cases). Genome-wide significant p-values are in bold. * List of genes in a LD region of *r*^2^=0.6 around the lead SNPs (±50 kb): **rs674094**: *SERPING1*(−233.0), *MIR130A*(−206.6), *YPEL4*(−197.9), *CLP1*(−186.0), *ZDHHC5*(−146.7), *MED19*(−135.5), *TMX2*(−106.9), *TMX2-CTNND1*(−28.7), *SELENOH*(−104.3), *BTBD18*(−96.1), *CTNND1*(−28.3); **rs9840050**: *SLC26A6*(−410.4), *MIR6824*(−412.2), *CELSR3*(−383.0), *MIR4793*(−401.6), *LINC02585*(−376.7), *NCKIPSD*(−360.0), *IP6K2*(−328.6), *PRKAR2A*(−198.0), *PRKAR2A-AS1*(−193.9), *SLC25A20*(−147.0), *ARIH2OS*(−126.5), *ARIH2*(−59.5), *P4HTM*(−38.7), *WDR6*(−29.9), *DALRD3*(−24.8), *MIR425*(−25.6), *NDUFAF3*(−22.4), *MIR191*(−25.2), *IMPDH2*(−16.5), *QRICH1*(0.0), *QARS*(0.0), *MIR6890*(0.0), *USP19*(0.0), *LAMB2*(0.0), *LAMB2P1*(7.0), *CCDC71*(16.7), *KLHDC8B*(25.7), *C3orf84*(31.8), *CCDC36*(52.5), *C3orf62*(122.7), *MIR4271*(128.2), *USP4*(131.3); **rs2564930**: *NEK4*(−162.4), *ITIH1*(−141.2), *ITIH3*(−124.3), *ITIH4*(−102.6), *ITIH4-AS1*(−108.0), *MUSTN1*(−98.3), *STIMATE-MUSTN1*(−35.7), *STIMATE* (−35.7), *MIR8064*(−86.7), *SFMBT1*(0.0), *RFT1*(55.2), *PRKCD*(127.9); **rs9265969**: *HCG20*(−505.7), *LINC00243*(−467.3), *LINC02570*(−449.8), *DDR1*(−397.8), *MIR4640*(−407.0), *GTF2H4*(−383.8), *VARS2*(−371.5), *SFTA2*(−365.8), *MUCL3*(−343.7), *HCG21*(−343.1), *MUC21*(−308.0), *MUC22*(−262.5), *HCG22*(−238.1), *C6orf15*(−185.4), *PSORS1C1*(−157.8), *CDSN*(−177.5), *PSORS1C2*(−158.9), *CCHCR1*(−139.7), *TCF19*(−133.7), *POU5F1*(−127.2), *PSORS1C3*(−111.6), *HCG27* (−94.0), *HLA-C*(−25.8), *LINC02571*(0.0), *HLA-B*(0.0), *MIR6891*(0.0), *MICA-AS1*(0.0), *MICA*(2.8), *LINC01149*(43.7), *HCP5*(65.2), *HCG26*(73.3), *MICB-DT* (82.0), *MICB*(97.0), *MCCD1*(131.0), *ATP6V1G2-DDX39B*(132.3), *DDX39B*(132.3), *SNORD117* (138.4), *SNORD84*(143.2), *DDX39B-AS1*(144.4), *ATP6V1G2*(146.5), *NFKBIL1*(148.9), *LOC100287329*(161.6), *LTA*(174.2), *TNF* (177.6), *LTB*(182.6), *LST1*(188.3), *NCR3*(191.0), *AIF1*(217.3), *PRRC2A*(222.8), *SNORA38*(225.1), *MIR6832*(235.9), *BAG6*(241.1), *APOM*(254.5), *C6orf47* (260.4), *GPANK1*(263.3), *CSNK2B*(267.9), *LY6G5B*(273.0), *LY6G5C*(278.8), *ABHD16A*(289.0), *MIR4646*(303.1), *LY6G6F* (308.9), *LY6G6F-LY6G6D*(308.9), *LY6G6E* (314.0), *LY6G6D*(317.4), *LY6G6C*(320.7), *MPIG6B*(325.4), *DDAH2*(329.1), *CLIC1*(332.7), *MSH5*(342.0), *MSH5-SAPCD1*(342.0), *SAPCD1*(365.1), *SAPCD1-AS1*(366.2), *VWA7* (367.7), *VARS*(379.6), *LSM2*(399.5), *HSPA1L*(411.7), *HSPA1A*(417.6), *HSPA1B*(429.8), *SNHG32*(437.0), *SNORD48*(437.3), *SNORD52*(439.1), *NEU1*(459.7), *SLC44A4*(465.3), *EHMT2*(481.8), *C2*(499.9), *ZBTB12*(501.7), *C2-AS1*(536.5), *CFB*(548.0), *NELFE* (554.2), *MIR1236*(558.9), *SKIV2L*(561.2), *DXO*(571.9), *STK19*(573.2), *C4A*(584.1), *C4B*(584.1), *C4B_2*(584.1), *LOC110384692*(584.1), *CYP21A1P*(607.7), *TNXA*(610.5), *TNXB*(610.5), *CYP21A2*(640.4), *ATF6B*(717.3), *FKBPL*(730.8); **rs4702**: *FURIN*(0.0), *FES*(0.0), *MAN2A2*(0.0), *HDDC3*(0.0), *UNC45A*(0.0); **rs3899258**: *PARP8*(−64.2), *LINC02106*(0.0); **rs2836950**: *PSMG1*(0.0), *BRWD1*(0.0), *BRWD1-AS2*(31.4), *BRWD1-AS1*(33.2), *HMGN1*(59.8), *GET1*(97.7), *WRB-SH3BGR*(97.8); **rs4460629**: *DCST1*(−61.9), *DCST1-AS1*(−48.9), *ADAM15*(−50.1), *EFNA4*(−43.3), *EFNA3*(−25.3), *EFNA1*(0.0), *SLC50A1*(0.0), *DPM3*(0.0), *KRT-CAP2*(0.0), *TRIM46*(0.0), *MUC1*(0.0), *MIR92B*(0.0), *THBS3*(0.0), *MTX1*(0.0), *GBAP1*(0.0), *GBA*(18.9), *FAM189B*(31.7), *SCAMP3*(40.4)

| SNP | CHR | BP | A1/A2 | Meta |  |  |  | freeze1+NORDiC |  |  | iPSYCH |  |  | 23andMe |  |  | Genes |
| --- | --- | --- | --- | --- | --- | --- | --- | --- | --- | --- | --- | --- | --- | --- | --- | --- | --- |
|  |  |  |  | FRQ cases | FRQ controls | log(OR) | P | FRQ cases | log(OR) | P | FRQ cases | log(OR) | P | FRQ cases | log(OR) | P |  |
| rs10877425 | 12 | 60751330 | G/A | 0.514 | 0.518 | -0.051 | **1.14E-10** | 0.508 | -0.044 | 0.128 | 0.481 | -0.086 | 0.006 | 0.518 | -0.049 | **9.16E-09** | - |
| rs674094 | 11 | 57665336 | C/A | 0.701 | 0.705 | -0.055 | **1.39E-10** | 0.691 | -0.099 | 0.001 | 0.674 | -0.122 | 2.44E-04 | 0.705 | -0.046 | 8.06E-07 | many (11) |
| rs11953498 | 5 | 45240783 | G/C | 0.18 | 0.184 | -0.065 | **3.36E-10** | 0.172 | -0.077 | 0.038 | 0.148 | -0.064 | 0.138 | 0.184 | -0.063 | **6.87E-09** | HCN1(0.0) |
| rs9840050 | 3 | 49133310 | G/A | 0.655 | 0.66 | -0.05 | **6.09E-10** | 0.639 | -0.04 | 0.171 | 0.624 | -0.064 | 0.046 | 0.66 | -0.05 | **1.12E-08** | CELSR3; many (31) |
| rs2564930 | 3 | 53017307 | T/C | 0.338 | 0.341 | -0.051 | **7.45E-10** | 0.317 | -0.143 | 1.98E-06 | 0.33 | -0.063 | 0.057 | 0.341 | -0.042 | 2.33E-06 | many (12) |
| rs9265969 | 6 | 31315706 | G/A | 0.874 | 0.874 | 0.072 | **2.34E-09** | 0.881 | 0.082 | 0.057 | 0.86 | 0.077 | 0.079 | 0.874 | 0.07 | 5.57E-08 | many (103) |
| rs6660196 | 1 | 168874798 | T/G | 0.638 | 0.637 | 0.048 | **3.26E-09** | 0.651 | 0.071 | 0.017 | 0.623 | 0.024 | 0.456 | 0.637 | 0.048 | **4.32E-08** | LINC00970(0.0) |
| rs4702 | 15 | 91426560 | G/A | 0.456 | 0.456 | 0.045 | **5.82E-09** | 0.458 | 0.046 | 0.109 | 0.451 | 0.067 | 0.04 | 0.456 | 0.044 | 1.56E-07 | many (5) |
| rs2198140 | 8 | 35039646 | T/C | 0.493 | 0.494 | -0.046 | **6.39E-09** | 0.483 | -0.045 | 0.113 | 0.497 | -0.046 | 0.137 | 0.494 | -0.046 | 7.24E-08 | UNC5D(3.3) |
| rs3899258 | 5 | 50256551 | G/A | 0.779 | 0.777 | 0.055 | **6.90E-09** | 0.788 | 0.055 | 0.113 | 0.781 | 0.008 | 0.84 | 0.777 | 0.059 | **8.84E-09** | many (2) |
| rs66818976 | 5 | 90900362 | G/A | 0.28 | 0.284 | -0.05 | **1.45E-08** | 0.261 | -0.092 | 0.005 | 0.275 | -0.03 | 0.396 | 0.284 | -0.048 | 4.25E-07 | - |
| rs2836950 | 21 | 40604429 | G/C | 0.361 | 0.361 | -0.046 | **2.40E-08** | 0.362 | -0.008 | 0.795 | 0.363 | -0.039 | 0.226 | 0.360 | -0.05 | **1.88E-08** | many (7) |
| rs35518360 | 4 | 103146890 | T/A | 0.0874 | 0.0899 | 0.076 | **3.78E-08** | 0.084 | 0.115 | 0.035 | 0.059 | 0.109 | 0.109 | 0.091 | 0.071 | 1.13E-06 | SLC39A8(0.0) |
| rs4460629 | 1 | 155135335 | T/C | 0.526 | 0.526 | 0.043 | **3.96E-08** | 0.54 | 0.025 | 0.375 | 0.5 | 0.036 | 0.239 | 0.526 | 0.045 | 9.28E-08 | many (18) |
| rs1922782 | 2 | 58916336 | T/C | 0.427 | 0.426 | 0.043 | **4.89E-08** | 0.44 | 0.098 | 5.8E-04 | 0.419 | 0.069 | 0.028 | 0.426 | 0.036 | 2.09E-05 | LINC01122(0.0) |

## Supplementary

### CONSORTIA

#### Members of the Nordic OCD & Related Disorders Consortium (NORDiC)

Julia Boberg, Long Long Chen, James J. Crowley, Elles de Schipper, Diana R. Djurfeldt, Jan Haavik, Kristen Hagen, Matthew W. Halvorsen, Bjarne Hansen, Kira D. Höffler, Anna K. Kähler, Elinor K. Karlsson, Gerd Kvale, Paul Lichtenstein, Kerstin Lindblad-Toh, Manuel Mattheisen, David Mataix-Cols, Kathleen Morrill, Hyun Ji Noh, Christian Rück, Stian Solem, Nora I. Strom, Tetyana Zayats

#### 23andMe Research Team

The following members of the 23andMe Research Team contributed to this study: Stella Aslibekyan, Adam Auton, Elizabeth Babalola, Robert K. Bell, Jessica Bielenberg, Katarzyna Bryc, Emily Bullis, Daniella Coker, Gabriel Cuellar Partida, Devika Dhamija, Sayantan Das, Sarah L. Elson, Teresa Filshtein, Kipper Fletez-Brant, Pierre Fontanillas, Will Freyman, Pooja M. Gandhi, Karl Heilbron, Barry Hicks, David A. Hinds, Ethan M. Jewett, Yunxuan Jiang, Katelyn Kukar, Keng-Han Lin, Maya Lowe, Jey C. McCreight, Matthew H. McIntyre, Steven J. Micheletti, Meghan E. Moreno, Joanna L. Mountain, Priyanka Nandakumar, Elizabeth S. Noblin, Jared O’Connell, Aaron A. Petrakovitz, G. David Poznik, Morgan Schumacher, Anjali J. Shastri, Janie F. Shelton, Jingchunzi Shi, Suyash Shringarpure, Vinh Tran, Joyce Y. Tung, Xin Wang, Wei Wang, Catherine H. Weldon, Peter Wilton, Alejandro Hernandez, Corinna Wong, Christophe Toukam Tchakouté.

#### PGC TS/OCD working group

Nora I. Strom^1,2,3,4^, Dongmei Yu^5,6^, Zachary F. Gerring^7^, Matthew W. Halvorsen^8^, Abdel Abdellaoui^9^, Cristina Rodriguez-Fontenla^10^, Julia M. Sealock^11^, Tim Bigdeli^12^, Jonathan R. I. Coleman^13,14^, Behrang Mahjani^15,16^, Jackson G. Thorp^17^, Katharina Bey^18^, Christie L. Burton^19^, Jurjen J. Luykx^20,21^, Gwyneth Zai^22,23^, Kathleen D. Askland^24^, CristinaBarlassina^25^, Judith Becker Nissen^26,27^, Laura Bellodi^28^, O. Joseph Bienvenu^29^, Donald Black^30^, Michael Bloch^31^, Julia Boberg^32^, Rosa Bosch^33^, Michael Breen^15,34,35^, Brian P. Brennan^36^, Helena Brentani^37^, Joseph D. Buxbaum^15^, JonasBybjerg-Grauholm^38^, Enda M. Byrne^39,40^, Beatriz Camarena^41^, Adrian Camarena^42^, Carolina Cappi^15,37^, Angel Carracedo^43^, Miguel Casas^44,45^, Maria C. Cavallini^46^, Valentina Ciullo^47^, Edwin H. Cook^48^, Vladimir Coric^31^, Bernadette A. Cullen^29^, Elles J. De Schipper^3^, Bernie Devlin^49^, Srdjan Djurovic^50,51^, Jason A. Elias^36^, Lauren Erdman^52^, Xavier Estivil^53^, Martha J.Falkenstein^54^, Bengt T. Fundin^16^, Maiken E. Gabrielsen^55^, Fernando S. Goes^29^, Marco A. Grados^29^, Jakob Grove^56,57^, Wei Guo^58,59^, Jan Haavik^60,61^, Kristen Hagen^62,63^, Alexandra Havdahl^64,65,66^, Ana G. Hounie^37^, Donald Hucks^11,67^, ChristinaHultman^16^, Magdalena Janecka^15,34^, Michael Jenike^68^, Elinor K. Karlsson^69,70^, Julia Klawohn^1^, Lambertus Klei^71^, JaniceKrasnow^72^, Kristi Krebs^73^, Jason Krompinger^36^, Nuria Lanzagorta^74^, Fabio Macciardi^75^, Brion Maher^76^, Evonne McArthur^11^, Nathaniel McGregor^77^, Nicole C. McLaughlin^78^, Sandra Meier^79^, Euripedes C. Miguel^37^, Maureen Mulhern^15,34^, Paul S.Nestadt^29^, Erika L. Nurmi^80^, Kevin S. O’Connell^81,82^, Lisa Osiecki^5,83^, Teemu Palviainen^84^, Fabrizio Piras^47^, Federica Piras^47^, Ann E. Pulver^29^, Raquel Rabionet^53^, Alfredo Ramirez^85,86,87,88^, Scott Rauch^54^, Abraham Reichenberg^89^, Jennifer Reichert^15,34^, Mark A. Riddle^29^, Stephan Ripke^6,90,91^, Aline S. Sampaio^37,92^, Miriam A. Schiele^93^, Laura G. Sloofman^15^, Jan Smit^94^, Janet L.Sobell^95^, María Soler Artigas^96,97,98,99^, Laurent F. Thomas^100,101^, Homero Vallada^37,102^, Jeremy Veenstra-VanderWeele^103^, Nienke N. C. C. Vulink^9^, Christopher P. Walker^104^, Ying Wang^29^, Jens R. Wendland^105^, Bendik S. Winsvold^106,107,108^, YinYao^109^, Pino Alonso^110^, Götz Berberich^111^, Cynthia M. Bulik^16,112,113^, Danielle Cath^114,115^, Daniele Cusi^116^, RichardDelorme^117^, Damiaan Denys^118^, Valsamma Eapen^119^, Peter Falkai^120^, Thomas V. Fernandez^31^, Abby J. Fyer^121,122^, Daniel A.Geller^5,123^, Hans J. Grabe^124^, Benjamin D. Greenberg^77,78,125^, Gregory L. Hanna^126^, Ian M. Hickie^127^, David M. Hougaard^38,57^,Norbert Kathmann^1^, James Kennedy^23^, Liang Kung-Yee^128,129^, Mikael Landén^16,130^, Stéphanie Le Hellard^131,132^, Marion Leboyer^133^, Christine Lochner^134^, James T. McCracken^80^, Sarah E. Medland^7^, Preben B. Mortensen^57,135,136^, Benjamin Neale^83,137,138^, Humberto Nicolini^139,140^, Merete Nordentoft^141,142^, Michele Pato^143^, Carlos Pato^143^, David L. Pauls^144^, Nancy L.Pedersen^16^, John Piacentini^80^, Christopher Pittenger^145^, Danielle Posthuma^146^, Josep A Ramos-Quiroga^147,148,149,150^, StevenA. Rasmussen^78^, Kerry J. Ressler^36^, Margaret A. Richter^23,151^, Maria C. Rosário^152^, David R. Rosenberg^153^, Stephan Ruhrmann^85^, Jack F. Samuels^29^, Sven Sandin^15,16^, Paul Sandor^23^, Gianfranco Spalletta^47,154^, Dan J. Stein^155^, S. Evelyn Stewart^156^, Eric A. Storch^154^, Barbara E. Stranger^157,158^, Maurizio Turiel^159^, Thomas Werge^57,142,160,161^, Ole A.Andreassen^82,162^, Anders D. Børglum^56,57^, Susanne Walitza^163,164,165^, Bjarne K. A. Hansen^131,166^, Christian P. Rück^3^, Nicholas G. Martin^17^, Lili Milani^73^, Ole Mors^167^, Ted Reichborn-Kjennerud^64,162^, Marta Ribasés^97,168,169,170^, Gerd Kvale^131,166^, DavidMataix-Cols^3^, Katharina Domschke^93,171^, Edna Grünblatt^163,164,165^, Michael Wagner^18^, John-Anker Zwart^106,107,172^, Gerome Breen^13,14^, Gerald Nestadt^29^, Andres Metspalu^73^, Jaakko Kaprio^173^, Paul D. Arnold^174,175^, Dorothy E. Grice^15^, James A.Knowles^176^, Helga Ask^64^, Karin J. H. Verweij^9^, Lea K. Davis^67^, Dirk J. A. Smit^177^, James J. Crowley^3,8,112^, Carol A. Mathews^178^, Eske M. Derks^17^, Jeremiah M. Scharf^5,6^, and Manuel Mattheisen2, 4, 180

^1^Department of Psychology, Humboldt-Universität zu Berlin, Berlin, Germany ^2^Institute of Psychiatric Phenomics and Genomics (IPPG), University Hospital of Munich, Munich, Germany ^3^Department of Clinical Neuroscience, Karolinska Institutet, Stockholm, Sweden ^4^Department of Biomedicine, Aarhus University, Aarhus, Denmark ^5^Department of Psychiatry, Massachusetts General Hospital, Boston, MA, USA ^6^Stanley Center for Psychiatric Research, Broad Institute of MIT and Harvard, Cambridge, MA, USA ^7^Department of Mental Health, QIMR Berghofer Medical Research Institute, Brisbane, QLD, Australia ^8^Department of Genetics, University of North Carolina At Chapel Hill, Chapel Hill, NC, USA ^9^Department of Psychiatry, Amsterdam UMC, University of Amsterdam, Amsterdam, The Netherlands ^10^CIMUS (centre for Research In Molecular Medicine and Chronic Diseases), University of Santiago De Compostela, Santiago De Compostela, A Coruña, Spain ^11^Vanderbilt Genetics Institute, Vanderbilt University Medical Center, Nashville, TN, USA ^12^Departments of Psychiatry and Behavioral Sciences, SUNY Downstate Health Sciences University, Brooklyn, NY, USA ^13^Social, Genetic and Developmental Psychiatry Centre, King’s College London, London, United Kingdom ^14^Departments of South London and Maudsley NHS Trust, NIHR Maudsley Biomedical Research Centre, London, United Kingdom ^15^Department of Psychiatry, Icahn School of Medicine At Mount Sinai, New York, NY, USA ^16^Departments of Medical Epidemiology and Biostatistics, Karolinska Institutet, Stockholm, Sweden ^17^Departments of Genetics and Computational Biology, QIMR Berghofer Medical Research Institute, Brisbane, QLD, Australia ^18^Departments of Psychiatry and Psychotherapy, University Hospital Bonn, Bonn, Germany ^19^Departments of Neurosciences and Mental Health, Hospital for Sick Children, Toronto, ON, Canada ^20^Department of Psychiatry, University Medical Center Utrecht, Utrecht, The Netherlands ^21^Second Opinion Outpatient Clinic, Ggnet, Warnsveld, The Netherlands ^22^Molecular Brain Science Department, Campbell Family Mental Health Research Institute, Centre for Addiction and Mental Health, Toronto, ON, Canada ^23^Department of Psychiatry, University of Toronto, Toronto, ON, Canada ^24^Waypoint Research Institute AND Outpatient Assessment and Treatment Services„ Waypoint Centre for Mental Health Care, Penetanguishene, ON, Canada ^25^Department of Heath Sciences, University of Milano, Milano, Milano, Italy ^26^Departments of Child and Adolescent Psychiatry, Aarhus University Hospital, Psychiatry, Denmark, Aarhus University Hospital, Psychiatry, Aarhus, Denmark ^27^Institute of Clinical Medicine, Health, Aarhus University, Health, Aarhus University, Aarhus, Danmark ^28^Department of Neuropsychiatric Sciences, Università Vita-salute San Raffaele Milano Italy, Milano, Italy ^29^Departments of Psychiatry and Behavioral Sciences, Johns Hopkins University, Baltimore, MD, USA ^30^Departments of Roy J. and Lucille A. Carver College of Medicine, University of Iowa, Iowa City, IA, USA ^31^Child Study Center and Psychiatry, Yale University, New Haven, CT, USA ^32^Center for Psychiatric Research, Institution of Clinical Neuroscience, Stockholm, Sweden ^33^Department of MIND SCHOOLS, HOSPITAL SANT JOAN DE DEU, ESPLUGUES DE LLOBREGAT, BARCELONA, Spain ^34^Seaver Autism Center for Research and Treatment, Icahn School of Medicine At Mount Sinai, New York, NY, USA ^35^The Mindich Child Health and Development Institute, Icahn School of Medicine Mount Sinai, New York, NY, USA ^36^Department of Mclean Hospital, Harvard Medical School, Belmont, MA, USA ^37^Department of Psychiatry, Universidade De São Paulo, São Paulo, Brazil ^38^Department of Congenital Disorders, Statens Serum Institut, Copenhagen, Denmark ^39^Institute for Molecular Bioscience, University of Queensland, Brisbane, Queensland, Australia ^40^Child Health Research Centre, University of Queensland, Brisbane, QLD, Australia ^41^Department of Pharmacogenetics, Instituto Nacional De Psiquitria Ramon De La Fuente Muñiz, Mexico City, Mexico ^42^Department of Surgery, Duke University, Durham, NC, USA ^43^Genomics Group, University of Santiago De Compostela, Santiago De Compostela, A Coruña, Spain ^44^Programa MIND Escoles, Hospital Sant Joan De Déu, ESPLUGUES DE LLOBREGAT, Barcelona, Spain ^45^Department of Departamento De Psiquiatría Y Medicina Legal, Universitat Autònoma De Barcelona, Bellaterra, Barcelona, Spain ^46^Department of Psychiatry, Ospedale San Raffaele, Milano, Italy ^47^Laboratory of Neuropsychiatry, IRCCS Santa Lucia Foundation, Rome, Italy ^48^Department of Psychiatry, University of Illinois At Chicago, Chicago, IL, USA ^49^Department of Psychiatry, University of Pittsburgh School of Medicine, Pittsburgh, PA, USA ^50^Department of Medical Genetics, Oslo University Hospital, Oslo, Norway ^51^NORMENT, Clinical Science, University of Bergen, Bergen, Norway ^52^Departments of Genetics and Genome Biology, The Hospital for Sick Children, Toronto, ON, Canada ^53^Department of Genetics, University of Barcelona, Barcelona, Spain ^54^Department of Psychiatry, Mclean Hospital, Harvard Medical School, Belmont, MA, USA ^55^Departments of Public Health and Nursing, Norwegian University of Science and Technology, Trondheim, Norway ^56^Biomedicine and The iSEQ Center, Aarhus University, Aarhus, Denmark ^57^The Lundbeck Foundation Initiative for Integrative Psychiatric Research, Ipsych,, Denmark ^58^National Institute of Mental Health, National Institutes of Health, Bethesda, MD, USA ^59^Genetic Epidemiology Research Branch, National Institute of Mental Health, National Institutes of Health, Bethesda, MD, Bethesda, MD, USA ^60^Department of Biomedicine, University of Bergen, Bergen, Norway ^61^Department of Division of Psychiatry, Haukeland University Hospital, Bergen, Norway ^62^Department of Molde Hospital, Møre Og Romsdal Hospital Trust, Molde, Norway ^63^Department of Mental Health, Norwegian University of Science and Technology, Trondheim, Norway ^64^Department of Mental Disorders, Norwegian Institute of Public Health, Oslo, Norway ^65^Nic Waals Institute, Lovisenberg Diaconal Hospital, Oslo, Norway ^66^PROMENTA Research Center, Psychology, University of Oslo, Oslo, Norway ^67^Department of Medicine, Vanderbilt University Medical Center, Nashville, TN, USA ^68^Department of Psychiatry, Massachusetts General Hospital and Harvard Medical School, Boston, MA, USA ^69^Departments of Bioinformatics and Integrative Biology, University of Massachusetts Medical School, Worcester, MA, USA ^70^Department of Vertebrate Genomics, Broad Institute of MIT and Harvard, Cambridge, MA, USA ^71^Department of Psychiatry, University of Pittsburgh, Pittsburgh, PA, USA ^72^Department of Psychiatry, Johns Hopkins University, Baltimore, MD, USA ^73^Institute of Genomics, University of Tartu, Tartu, Estonia ^74^Carracci Medical Group, Mexico City, Mexico ^75^Departments of Psychiatry and Human Behavior, University of Californiairvine (UCI), Irvine, CA, USA ^76^Department of Mental Health, Johns Hopkins Bloomberg School of Public Health, Baltimore, MD, USA ^77^COBRE Center for Neuromodulation, Butler Hospital, Providence, RI, USA ^78^Departments of Psychiatry and Human Behavior, Alpert Medical School, Brown University, Providence, RI, USA ^79^Department of Psychiatry, Dalhousie University, Halifax, NS, Canada ^80^Departments of Psychiatry and Biobehavioral Sciences, University of California, Los Angeles, Los Angeles, CA, USA ^81^NORMENT Center of Excellence (coe), Institute of Clinical Medicine, University of Oslo, Oslo, Norway ^82^Departments of Division of Mental Health and Addiction, Oslo University Hospital, Oslo, Norway ^83^Psychiatric and Neurodevelopmental Genetics Unit, Harvard Medical School, Boston, MA, USA ^84^Institute for Molecular Medicine Finland - FIMM, University of Helsinki, Helsinki, Finland ^85^Departments of Psychiatry and Psychotherapy, University of Cologne, Cologne, Germany ^86^Departments of Neurodegenerative Diseases and Geriatric Psychiatry, University of Bonn, Bonn, Germany ^87^DZNE Bonn, German Center for Neurodegenerative Diseases (DZNE), Bonn, Germany ^88^Psychiatry and Glenn Biggs Institute for Alzheimer’s and Neurodegenerative Diseases, UT Health San Antonio, San Antonio, TX, USA ^89^Department of Mental Disorders, Norwegian Institute of Public Health, New York, NY, USA ^90^Department of Medicine, Massachusetts General Hospital and Harvard Medical School, Boston, MA, USA ^91^Departments of Psychaitry and Psychotherapy, Charité Universitätsmedizin, Berlin, Germany ^92^Department of University Health Careservices - SMURB, Federal University of Bahia, Salvador, Brazil ^93^Departments of Psychiatry and Psychotherapy, Medical Center - University of Freiburg, Freiburg, Germany ^94^Department of Psychiatry, Amsterdam UMC Location Vumc, Amsterdam, The Netherlands ^95^Departments of Psychiatry and The Behavioral Sciences, Keck School of Medicine of USC, Los Angeles, CA, USA ^96^Psychiatric Genetics Unit, Group of Psychiatry, Mental Health and Addiction, Vall D’hebron Research Institute (VHIR), Barcelona, Spain ^97^Department of Psychiatry, Hospital Universitari Vall D’hebron, Barcelona, Spain ^98^Department of Instituto De Salud Carlos III, Biomedical Network Research Centre On Mental Health (CIBERSAM), Madrid, Spain ^99^Department of Biology, Universitat De Barcelona, Barcelona, Spain ^100^Departments of Clinical and Molecular Medicine, Norwegian University of Science and Technology, Trondheim, Norway ^101^K. G. Jebsen Center for Genetic Epidemiology, Norwegian University of Science and Technology, Trondheim, Norway ^102^Departments of Molecular Medicine and Surgery, Karolinska Institutet, Stockholm, Sweden ^103^Departments of Psychiatry, Pediatrics, and Pharmacology, Vanderbilt University Medical Center, Nashville, TN, USA ^104^Department of Precision Medicine, City of Hope, Monrovia, CA, USA ^105^Laboratory of Clinical Science, NIMH Intramural Research Program, Bethesda, MD, USA ^106^Departments of Research, Innovation and Education, Oslo University Hospital, Oslo, Norway ^107^Departments of Medicine and Health Sciences, Norwegian University of Science and Technology, Trondheim, Norway ^108^Department of Neurology, Oslo University Hospital, Oslo, Norway ^109^Department of Computional Biology„ Fudan University., Fudan, PRC ^110^Department of Bellvitge University, University of Barcelona, Barcelona, Spain ^111^Psychosomatic Department, Windach Hospital of Neurobehavioural Research and Therapy, Windach, Germany ^112^Department of Psychiatry, University of North Carolina At Chapel Hill, Chapel Hill, NC, USA ^113^Department of Nutrition, University of North Carolina At Chapel Hill, Chapel Hill, NC, USA ^114^Departments of Rijksuniversiteit Groningen and Psychiatry, University Medical Center Groninge, Groningen, The Netherlands ^115^Department of Specialized Training, Drenthe Mental Health Care Institute,, ^116^Institute of Biomedical Technologies, Italian National Centre for Research, Segrate, Milano, Italy ^117^Child and Adolesccent Psycchiatry Department, APHP, Paris, France ^118^Department of Psychiatry, Institute of The Royalnetherlands Academy of Arts and Sciences (NIN-KNAW), Amsterdam, The Netherlands ^119^Department of Medicine, University of New South Wales, Randwick, NSW, Australia ^120^Departments of Psychiatry and Psychotherapy, University of Göttingen, Göttingen, Germany ^121^Department of Psychiatry, New York State Psychiatric Institute, New York, NY, USA ^122^Department of Psychiatry, Columbia University Medical Center, New York, NY, USA ^123^Department of Psychiatry, Harvard Medical School, Boston, MA, USA ^124^Departments of Psychiatry and Psychotherapy, University Medicine Greifswald, Greifswald, Greifswald ^125^RR&D Center for Neurorestoration and Neurotechnology, Providence VA Medical Center, Providence, RI ^126^Department of Psychiatry, University of Michigan, Ann Arbor, MI, USA ^127^Brain and Mind Center, The University of Sydney, Sydney, NSW, Australia ^128^Department of Biostatistics, Johns Hopkins Bloomberg School of Public Health, Baltimore, MD, USA ^129^Institute of Population Health Sciences, National Health Research Institutes,, Taiwan ^130^Institute of Neuroscience and Physiology, University of Gothenburg, Gothenburg, Sweden ^131^Bergen Center for Brain Plasticity, Haukeland University Hospital, Bergen, Norway ^132^Department of Clinical Science, University of Bergen, Bergen, Norway ^133^Departments of Addictology and Psychiatry, Univ Paris Est Créteil, AP-HP, Inserm, Paris, France ^134^Department of Psychiatry, Stellenbosch University, Stellenbosch, South Africa ^135^National Centre for Register-based Research, Aarhus University, Aarhus, Denmark ^136^Centre for Integrated Register-based Research, Aarhus University, Aarhus, Denmark ^137^Analytic and Translational Genetics Unit, Massachusetts General Hospital, Boston, MA, USA ^138^Program In Medical and Population Genetics, Broad Institute of Harvard and MIT, Cambridge, MA, USA ^139^Department of Psychiatric Genetics, Grupo Médico Carracci INMEGEN, Mexico City, México ^140^Department of Psychiatric Genetics, Instituto Nacional De Medicina Genómica, México City, Mexico ^141^CORE - Copenhagen Research Center for Mental Health, Copenhagen University Hospital, Copenhagen, Denmark ^142^Department of Clinical Medicine, University of Copenhagen, Copenhagen, Denmark ^143^Department of Psychiatry, Rutgers University, Piscataway, NJ, USA ^144^Department of Psychiatry, Harvard University, Boston, MA, USA ^145^Department of Psychiatry, Yale University, New Haven, CT, USA ^146^Department of Clinical Genetics, VU University Medical Center Amsterdam, Amsterdam, The Netherlands ^147^Department of Psychiatry, Hospital Universitari Vall D’hebron, Barcelona, Catalonia, Spain ^148^Group of Psychiatry, Mental Health and Addictions, Vall D’hebron Research Institute (VHIR), Barcelona, Spain ^149^Group 27, Biomedical Network Research Centre On Mental Health (CIBERSAM), Barcelona, Spain ^150^Departments of Psychiatry and Forensic Medicine, Universitat Autònoma De Barcelona, Barcelona, Spain ^151^Department of Psychiatry, Sunnybrook Health Sciences Centre,, ^152^Department of Psychiatry, Federal University of São Paulo (UNIFESP), São Paulo, Brazil ^153^Departments of Psychiatry and Behavioral Neurosciences, Wayne State University School of Medicine, Detroit, MI, USA ^154^Departments of Psychiatry and Behavioral Sciences, Baylor College of Medicine, Houston, TX, USA ^155^Psychiatry and Neuroscience Institute, University of Cape Town, Cape Town, Western Cape, South Africa ^156^Department of Psychiatry, University of British Columbia, Vancouver, BC, Canada ^157^Department of Pharmacology, Northwestern University Feinberg School of Medicine, Chicago, IL, USA ^158^Center for Genetic Medicine, Northwestern University Feinberg School of Medicine, Chicago, IL, USA ^159^Department of Cardiology, University of Milan, Milan, Italy ^160^Department of Mental Health Services, Copenhagen University Hospital, Copenhagen, Denmark ^161^GLOBE Institute, University of Copenhagen, Copenhagen, Denmark ^162^Institute of Clinical Medicine, University of Oslo, Oslo, Norway ^163^Departments of Child and Adolescent Psychiatry and Psychotherapy, University of Zurich, Zürich, Switzerland ^164^Neuroscience Center Zurich, University of Zurich and The ETH Zuric, Zurich, Switzerland ^165^Zurich Center for Integrative Human Physiology, University of Zurich, Zurich, Switzerland ^166^Department of Psychology, University of Bergen, Bergen, Norway ^167^Psychosis Reasearch Unit, Aarhus University Hospital - Psychiatry, 8200 Aarhus N, Denmark ^168^Psychiatric Genetics Unit, Vall D’hebron Research Institute (VHIR), Universitat Autònoma De Barcelona, Barcelona, Spain ^169^Biomedical Network Research Centre On Mental Health (CIBERSAM), Barcelona, Spain. 3 Biomedical Network Research Centre On Mental Health (CIBERSAM), Instituto De Salud Carlos III, Madrid, Spain ^170^Departments of Genetics, Microbiology and Statistics, University of Barcelona, Barcelona, Spain ^171^Center for Basics In Neuromodulation, University of Freiburg, Freiburg, Germany ^172^Department of Medicine, University of Oslo, Oslo, Norway ^173^Helsinki Institute of Life Science, University of Helsinki, Helsinki, Finland ^174^Department of Psychiatry, Cumming School of Medicine, University of Calgary, Calgary, AB, Canada ^175^Program In Genetics and Genome Biology, Hospital for Sick Children, Toronto, ON, Canada ^176^Department of Cell Biology, SUNY Downstate Health Sciences University, Brooklyn, NY, USA ^177^Department of Psychiatry, Amsterdam UMC Location AMC, Amsterdam, The Netherlands ^178^Psychiatry and Genetics Institute, Center for OCD, Anxiety and Related Disorders, University of Florida, Gainesville, FL, USA ^179^Department of Psychiatry, Center for Genomic Medicine, Massachusetts General Hospital, Harvard Medical School, Boston, MA, USA ^180^Departments of Psychiatry and Community Health and Epidemiology, Dalhousie University, Halifax, NS, Canada

### GWAS details for NORDiC-NOR & NORDiC-SWE

#### pre-GWAS Quality Control

##### NORDiC-SWE

We formed a case/control dataset by merging PLINK filesets for four different cohorts (one case-only, three control-only) using PLINK v1.90b3n. This merged fileset consisted of 647,335 variant calls across a total of 1107 cases and 3,500 controls.

We first identified a subset of samples that will be usable in the final GWAS based on a lack of cryptic relatedness patterns with other samples and likely european ancestry. In preparation for this we used PLINK v1.90b3n to extract common (MAF > 5%), and used the LD-pruned (–indep 50 5 2) subset of these variants for relatedness checks, implemented via PLINK’s ‘genome’ function. As part of relatedness QC we first removed a total of 48 samples with mean pi hat values observed with other samples >= 0.02, which we considered to be evidence of systemic issues with the sample including but not limited to overall high genotype missingness. Next we pruned instances of sample duplication, defined as pi hat >= 0.95. For a set of samples which based on these criteria are duplicates, we selected the one sample to keep based on which sample carried the lowest genotype missingness, as computed in PLINK. A total of 3 samples were removed in the duplicate pruning process. In the remaining samples we identified all pairs where a cryptic relatedness pattern (defined here as pi hat > 0.2) is present. To resolve these relatedness issues in a manner that minimizes the total number of samples removed from the cohort, we used an iterative strategy that for each round, 1) defined rel_max as the maximum number of cryptic relationships observed across single samples, 2) removed samples where the number of cryptic relationships equaled rel_max, 3) recomputes rel_max in the pruned sampleset. Once rel_max equals 1, then for each pairwise cryptic relationship, we selected the sample to keep based on which sample carried the lower overall genotype missingness, as computed in PLINK. A total of 102 samples that carried cryptic relatedness issues were marked for exclusion from the final GWAS, leaving behind 1074 cases and 3380 controls. To identify the subset of samples that are of likely European ancestry, we utilized PEDDY v0.4.3, which allows for PCA of input samples with 1000 genomes sample data and subsequent classification per sample of its most likely general ethnicity. While PEDDY was originally designed primarily for exome and whole genome sequence data, it should be sufficient for identification of and pruning of samples that clearly are not of European ancestry. We found that a total of 6,039 variants overlapped between the merged PLINK fileset and the 1000 genomes data included in PEDDY, and used these to conduct PCA and PEDDY’s classification algorithm. We kept all samples with an ancestry classification of EUR. From the relatedness and ethnicity classification steps we identified a total of 989 cases and 3200 controls suitable for the final NORDIC GWAS.

To prepare our merged cohort dataset for the GWAS pipeline, we pruned single variants from our relatedness-pruned dataset (1,074 cases, 3,380 controls) where there was suggestive evidence of technical biases or batch effects. For this we first collect cohort level allele frequency and genotype missingness values for the QC-passing subset of input samples, and look for instances where an input cohort has an extreme value which might be consistent with batch effects or technical problems. We defined variants as QC-failing if they met one of the following criteria: 1) maximum genotype missingness in a cohort > 0.02; 2) allele frequency <0.001 in at least one cohort; 3) max – min allele frequency > 0.1 across all four cohorts; 4) max – min allele frequency > 0.03 across all three control cohorts; 5) genome-wide significant in a control vs. control synthetic GWAS. A total of 154,791 variants met at least one of these criteria were excluded, leaving us with a final case/control dataset consisting of genotype calls across 492,554 variants across the 1074 cases and 3,380 controls.

##### NORDiC-NOR

The pre-GWAS QC applied to the NORDiC-NOR dataset (482 cases, 343 controls in the raw data) was nearly identical to that applied to NORDiC-SWE data after the merging of separate genotype data, consisting of pruning of cryptic relatedness, marking of samples that are of likely European ancestry and pruning of the dataset for single variants where there was suggestive evidence of technical biases or batch effects. In our cryptic relatedness QC step we identified 4 samples that had a mean pi_hat with other samples >= 0.1, 74 samples that had evidence of being a sample duplicate (pi_hat >= 0.95) and 8 samples from the remaining cohort with evidence of cryptic relatedness (pi_hat >= 0.2), leaving behind 404 cases and 335 controls. We found a total of 6263 variants overlapped between the merged PLINK fileset and the 1000 genomes data included in PEDDY, and identified a total of 368 cases and 315 controls with likely European ancestry. We performed variant-level QC on a PC-pruned subset of 340 cases and 307 controls, defining variants as failing if they met one of the following criteria : 1) maximum genotype missingness in a cohort > 0.02; 2) allele frequency of 0 in at least one cohort; 3) max – min allele frequency > 0.1. A total of 136,525 variants met at least one of these criteria were excluded, leaving us with a final case/control dataset consisting of genotype calls across 479,358 variants. We used calls across these variants in samples that had been pruned for relatedness issues (not including standard pairwise relatedness issues as ricopili can detect these) as input for the GWAS (407 cases, 340 controls).

#### QC and imputation implemented in Ricopili

We used the ricopili pipeline to run an automated round of pre-imputation QC on the genotype data from 492,554 QC-passing variants across the full unpruned datasets of NORDiC-SWE (1107 cases and 3500 controls) and 479,358 QC-passing variants across NORDIC-Norway (407 cases and 340 controls), and conducted imputation on the genotype data using the Haplotype Reference Consortium (HRC) reference panel. All subsets of the analysis involved ricopili were done using ricopili v2018_Dec_7.001. The pre-imputation ricopili QC step involved a series of hard filters on variant and sample level data, including removing variants with pre-sample pruning call rate < 0.95, samples with call rate < 0.98, FHET outside of +/-0.20, samples with discrepancies between reported and derived sex, and post sample-pruning variants that meet any of the following : 1) call rate < 0.98, 2) missing difference > 0.02, 3) invariant positions, 4) MAF > 0.01, 5) HWE p < 1e-6 in controls, and 6) HWE p < 1e-10 in cases. After the QC described we were left with 481,323 QC-passing variants across 1077 cases and 3486 controls in NORDiC-SWE, and 469,618 QC-passing variants across 404 cases and 340 controls in NORDIC-Norway.

After sample and variant pruning we next used ricopili’s impute_dirsub module to perform imputation using HRC genotypes as a reference panel. In our imputation run we used eagle v2.3.5 for pre-phasing, and minimac3 v2.0.1 for imputation. We derived 3 different imputed callsets from this process: 1) a set of high confidence imputed genotypes (2,771,425 in NORDiC-SWE, 3,043,464 in NORDIC-Norway), 2) a set of imputed best-guess genotypes with medium level accuracy (7,112,906 in NORDiC-SWE, 7,277,174 in NORDIC-Norway), and 3) a set of for variants where imputation accuracy is lowered in order to increase the total number of variants included in the imputation (9,021,638 in NORDiC-SWE, 8,964,589 in NORDIC-Norway).

## Supplementary Figures

**Fig. S1.**
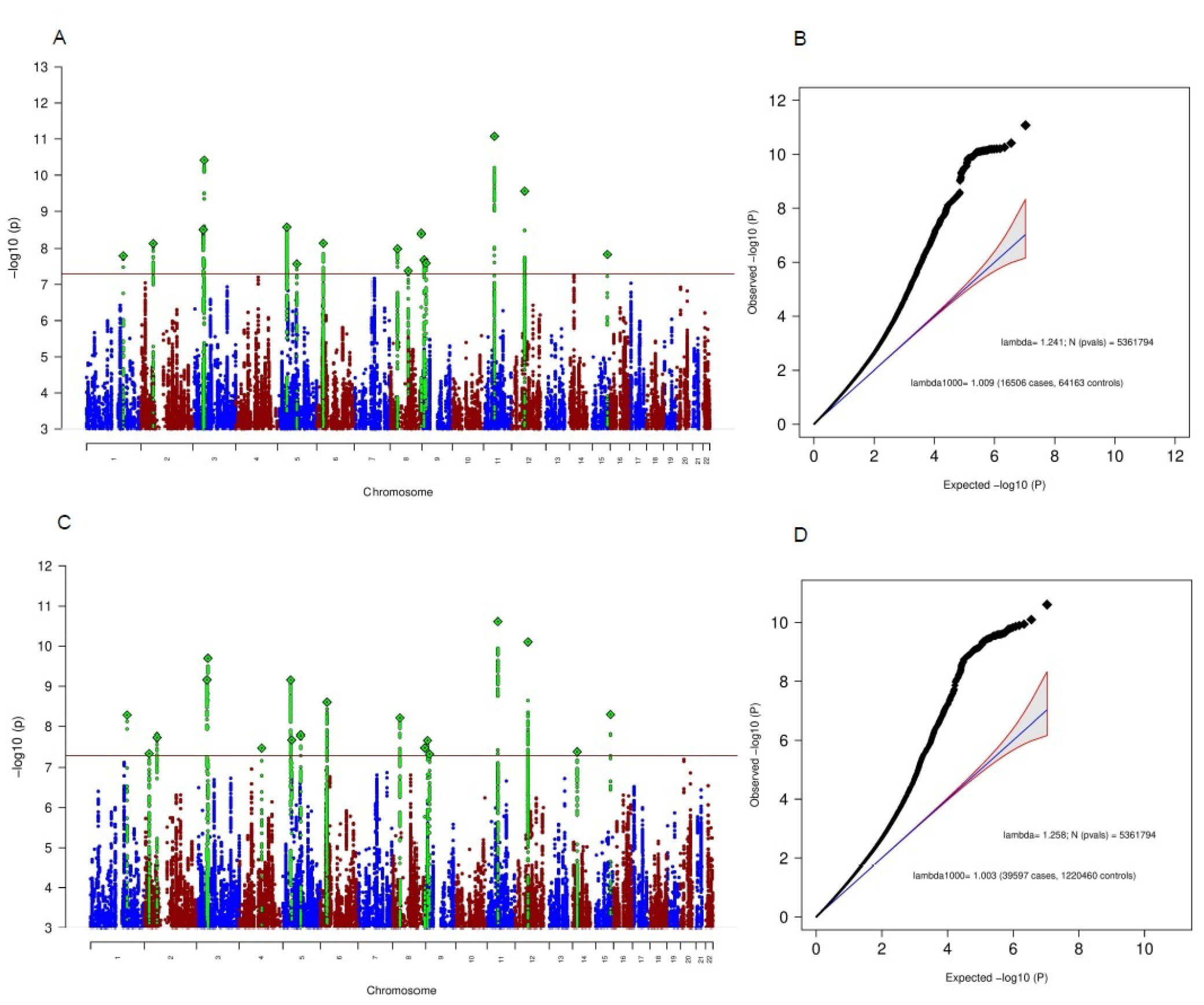
**Genome-wide association results for the MTAG OCD with cases ascertained for another disorder as the primary phenotype (iPSYCH; Manhattan plot in (A) and quantile-quantile plot in (B)) and for the MTAG OCD analysis with the biobank cases (23andMe) as the primary phenotype (Manhattan plot in (C) and quantile-quantile plot in (D)). The x-axis in (A) and (C) shows the position in the genome (chromosomes 1-22), while the y-axis represents –log10(p) values for the association of variants with OCD, The horizontal red line represents the threshold for genome-wide significance. Each dot represents one SNP that was tested in the GWAS, with green diamonds indicating the lead SNP in regions harboring a genome-wide significant SNP. In (B) and (D) the expected -log10(p) under the null is plotted against the observed -log10(p), qq-plots (B) and (D) belong to Manhattan lots (A) and (C), respectively. The shading indicates 95% confidence region under the null. Lambda indicates the genomic inflation factor**.

**Fig. S2.**
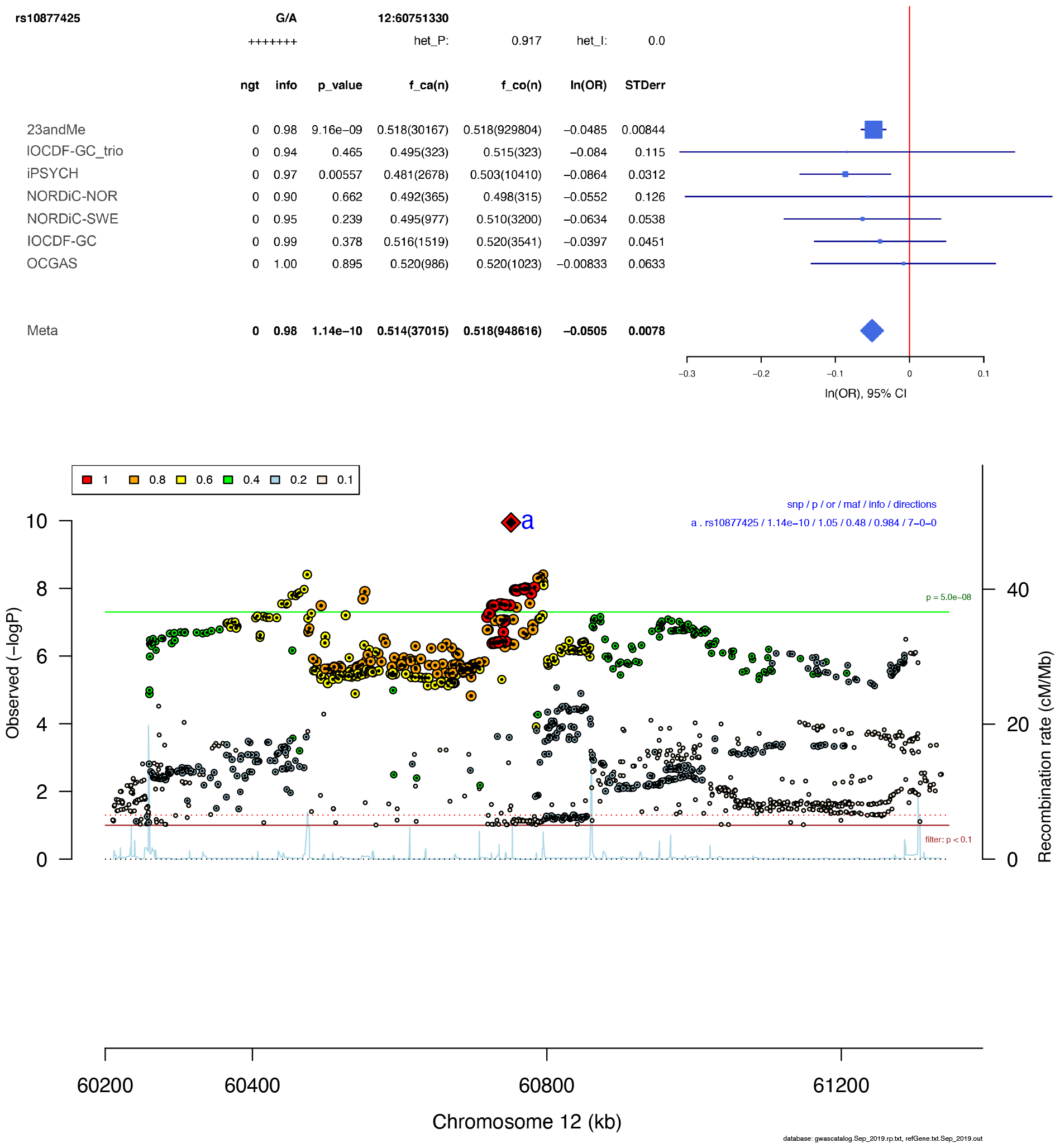
Forest plot (top) and regional association plot (bottom) of SNP rs10877425. Forest plot: Shows the imputation score (info), p-value (p_value), A1 allele frequency in cases and number of cases (f_ca(n)), A1 allele frequency in controls and number of controls (f_co(n)), effect of the association (ln(OR)), and standard error of the effect (STDerr) for each individual dataset and the over-all meta-analysis. On the right side, effects (ln(OR)) and 95% confidence intervals (CI) are plotted. At the top, the direction of effect for each study is shown (+ for positive effect of A1, - for negative effect of A1), and results of a test of heterogeneity (het_P and het_I) of effect across the individual datasets are displayed. Regional association plot: The –log10(P) of SNPs in the OCD meta-analysis GWAS is shown on the left y axis. The recombination rates expressed in centimorgans (cM) per Mb (Megabase) (blue line) are shown on the right y axis. Position in Mb is on the x axis. Only the SNPs with association p-value less than 0.1 were plotted. The lead SNP in the region is shown as a red diamond. Colour coding indicates LD to the lead SNP.

**Fig. S3.**
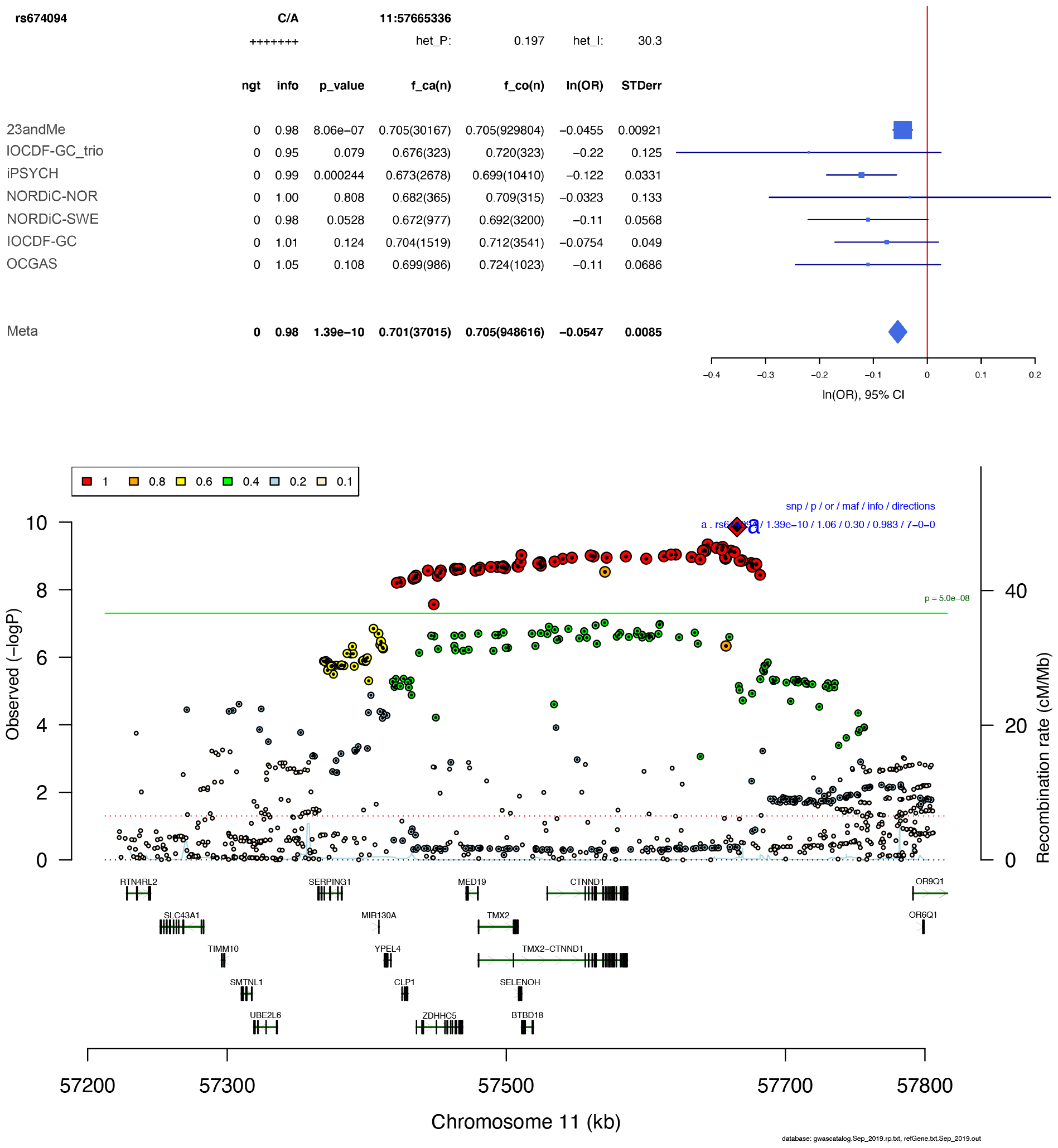
Forest plot (top) and regional association plot (bottom) of SNP rs674094. Forest plot: Shows the imputation score (info), p-value (p_value), A1 allele frequency in cases and number of cases (f_ca(n)), A1 allele frequency in controls and number of controls (f_co(n)), effect of the association (ln(OR)), and standard error of the effect (STDerr) for each individual dataset and the over-all meta-analysis. On the right side, effects (ln(OR)) and 95% confidence intervals (CI) are plotted. At the top, the direction of effect for each study is shown (+ for positive effect of A1, - for negative effect of A1), and results of a test of heterogeneity (het_P and het_I) of effect across the individual datasets re displayed. Regional association plot: The –log10(P) of SNPs in the OCD meta-analysis GWAS is shown on the left y axis. The recombination rates expressed in centimorgans (cM) per Mb (Megabase) (blue line) are shown on the right y axis. Position in Mb is on the x axis. Only the SNPs with association p-value less than 0.1 were plotted. The lead SNP in the region is shown as a red diamond. Colour coding indicates LD to the lead SNP.

**Fig. S4.**
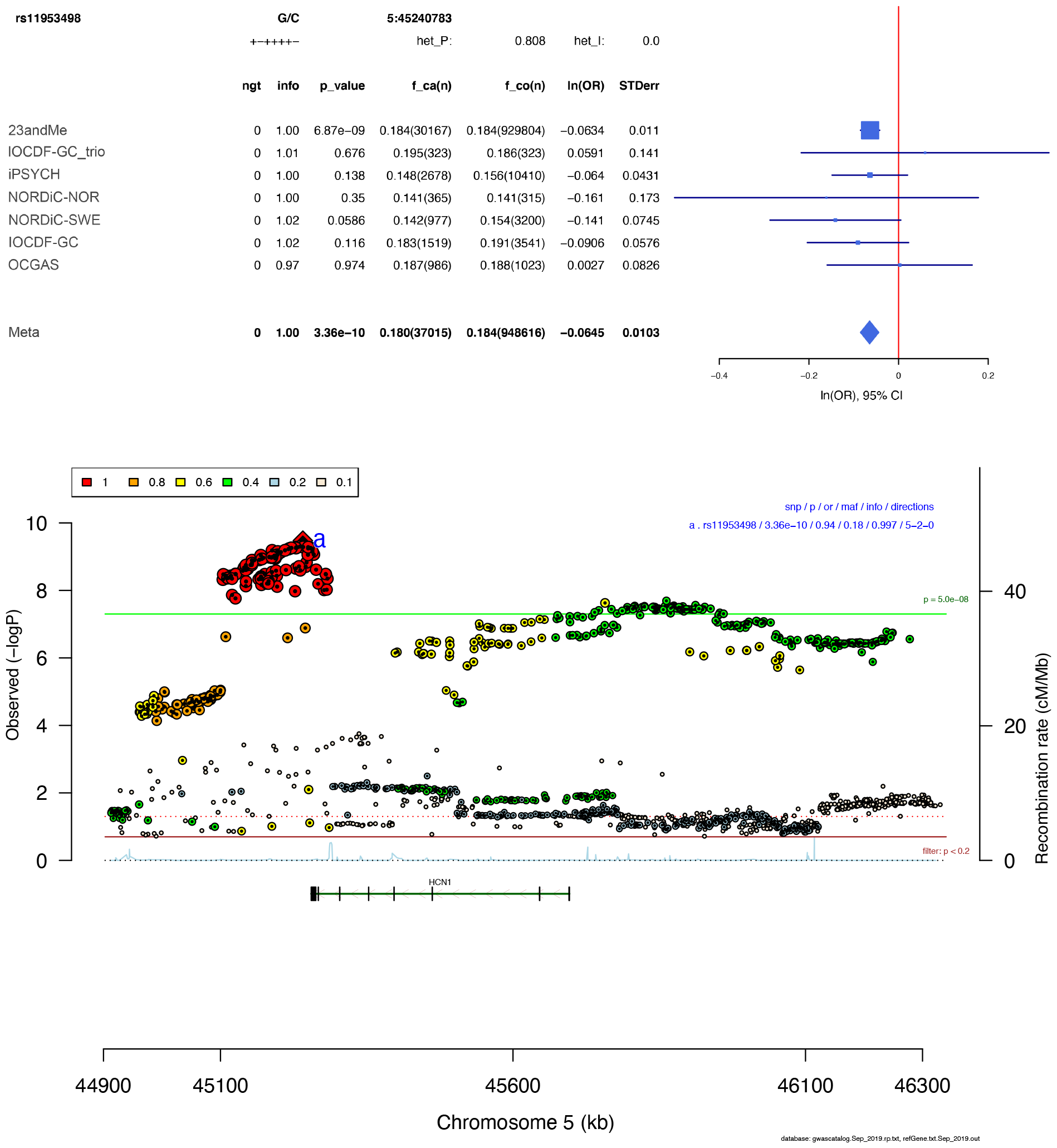
Forest plot (top) and regional association plot (bottom) of SNP rs11953498. Forest plot: Shows the imputation score (info), p-value (p_value), A1 allele frequency in cases and number of cases (f_ca(n)), A1 allele frequency in controls and number of controls (f_co(n)), effect of the association (ln(OR)), and standard error of the effect (STDerr) for each individual dataset and the over-all meta-analysis. On the right side, effects (ln(OR)) and 95% confidence intervals (CI) are plotted. At the top, the direction of effect for each study is shown (+ for positive effect of A1, - for negative effect of A1), and results of a test of heterogeneity (het_P and het_I) of effect across the individual datasets re displayed. Regional association plot: The –log10(P) of SNPs in the OCD meta-analysis GWAS is shown on the left y axis. The recombination rates expressed in centimorgans (cM) per Mb (Megabase) (blue line) are shown on the right y axis. Position in Mb is on the x axis. Only the SNPs with association p-value less than 0.1 were plotted. The lead SNP in the region is shown as a red diamond. Colour coding indicates LD to the lead SNP.

**Fig. S5.**
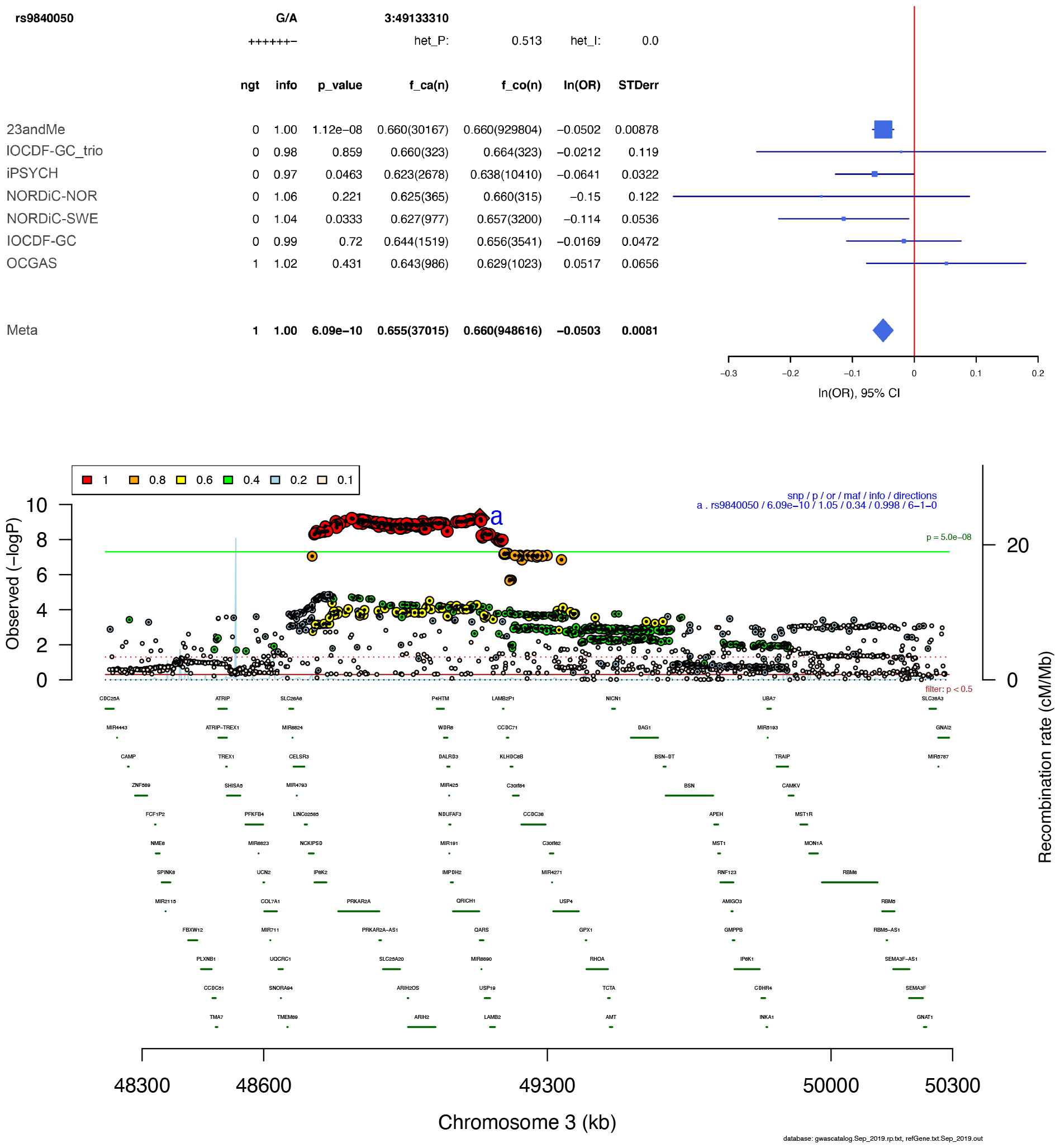
Forest plot (top) and regional association plot (bottom) of SNP rs9840050. Forest plot: Shows the imputation score (info), p-value (p_value), A1 allele frequency in cases and number of cases (f_ca(n)), A1 allele frequency in controls and number of controls (f_co(n)), effect of the association (ln(OR)), and standard error of the effect (STDerr) for each individual dataset and the over-all meta-analysis. On the right side, effects (ln(OR)) and 95% confidence intervals (CI) are plotted. At the top, the direction of effect for each study is shown (+ for positive effect of A1, - for negative effect of A1), and results of a test of heterogeneity (het_P and het_I) of effect across the individual datasets re displayed. Regional association plot: The –log10(P) of SNPs in the OCD meta-analysis GWAS is shown on the left y axis. The recombination rates expressed in centimorgans (cM) per Mb (Megabase) (blue line) are shown on the right y axis. Position in Mb is on the x axis. Only the SNPs with association p-value less than 0.1 were plotted. The lead SNP in the region is shown as a red diamond. Colour coding indicates LD to the lead SNP.

**Fig. S6.**
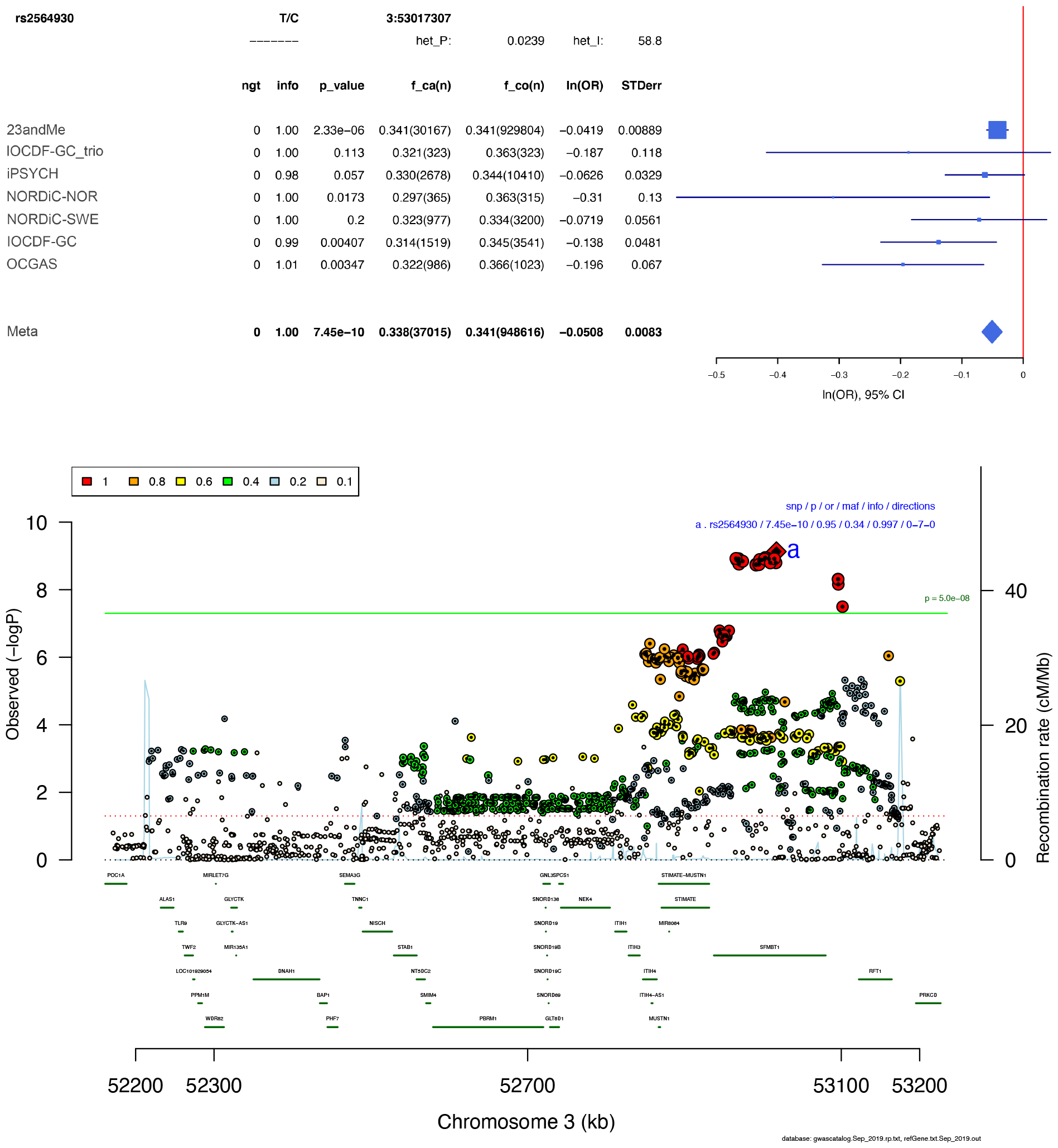
Forest plot (top) and regional association plot (bottom) of SNP rs2564930. Forest plot: Shows the imputation score (info), p-value (p_value), A1 allele frequency in cases and number of cases (f_ca(n)), A1 allele frequency in controls and number of controls (f_co(n)), effect of the association (ln(OR)), and standard error of the effect (STDerr) for each individual dataset and the over-all meta-analysis. On the right side, effects (ln(OR)) and 95% confidence intervals (CI) are plotted. At the top, the direction of effect for each study is shown (+ for positive effect of A1, - for negative effect of A1), and results of a test of heterogeneity (het_P and het_I) of effect across the individual datasets re displayed. Regional association plot: The –log10(P) of SNPs in the OCD meta-analysis GWAS is shown on the left y axis. The recombination rates expressed in centimorgans (cM) per Mb (Megabase) (blue line) are shown on the right y axis. Position in Mb is on the x axis. Only the SNPs with association p-value less than 0.1 were plotted. The lead SNP in the region is shown as a red diamond. Colour coding indicates LD to the lead SNP.

**Fig. S7.**
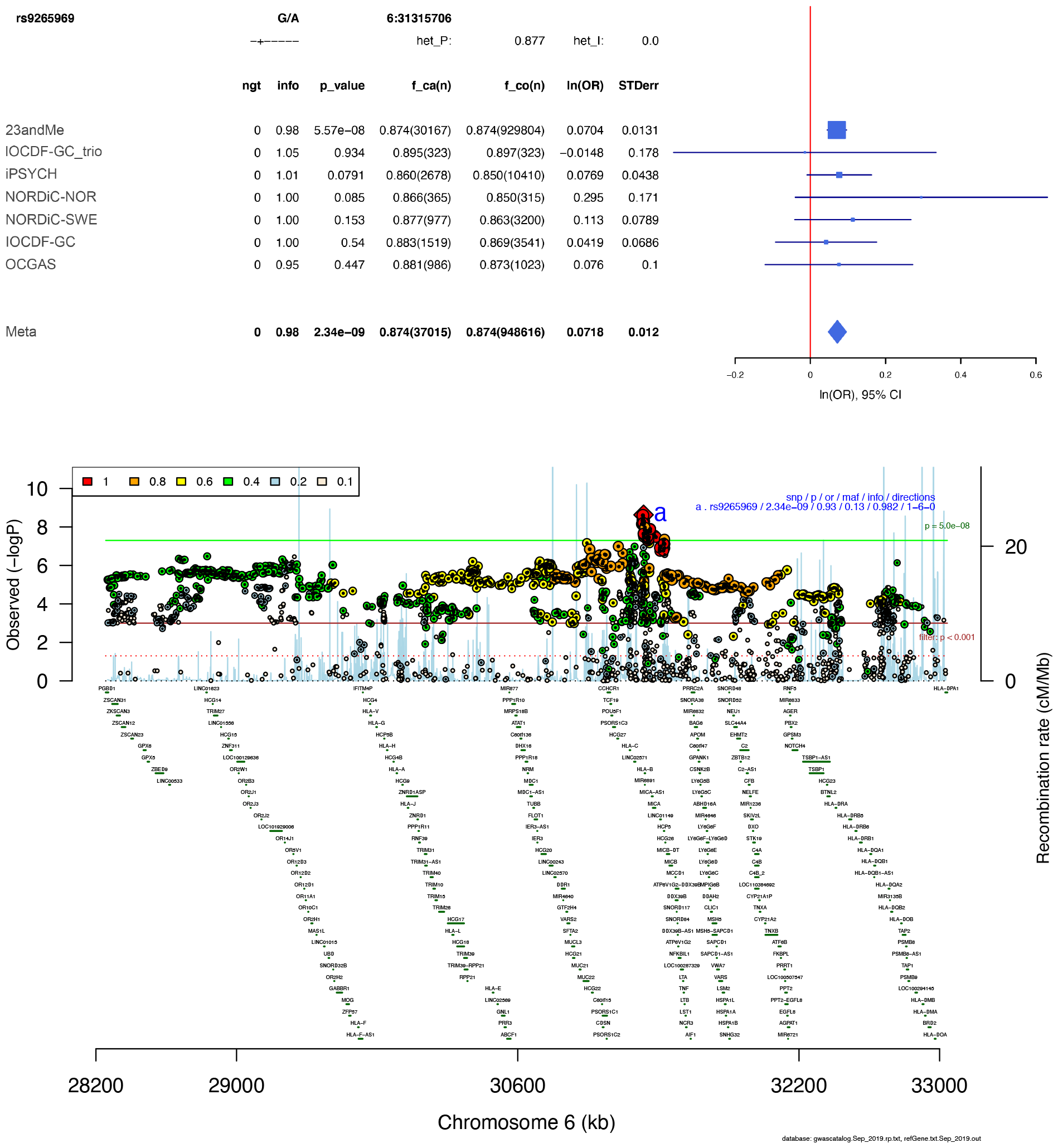
Forest plot (top) and regional association plot (bottom) of SNP rs9265969. Forest plot: Shows the imputation score (info), p-value (p_value), A1 allele frequency in cases and number of cases (f_ca(n)), A1 allele frequency in controls and number of controls (f_co(n)), effect of the association (ln(OR)), and standard error of the effect (STDerr) for each individual dataset and the over-all meta-analysis. On the right side, effects (ln(OR)) and 95% confidence intervals (CI) are plotted. At the top, the direction of effect for each study is shown (+ for positive effect of A1, - for negative effect of A1), and results of a test of heterogeneity (het_P and het_I) of effect across the individual datasets re displayed. Regional association plot: The –log10(P) of SNPs in the OCD meta-analysis GWAS is shown on the left y axis. The recombination rates expressed in centimorgans (cM) per Mb (Megabase) (blue line) are shown on the right y axis. Position in Mb is on the x axis. Only the SNPs with association p-value less than 0.1 were plotted. The lead SNP in the region is shown as a red diamond. Colour coding indicates LD to the lead SNP.

**Fig. S8.**
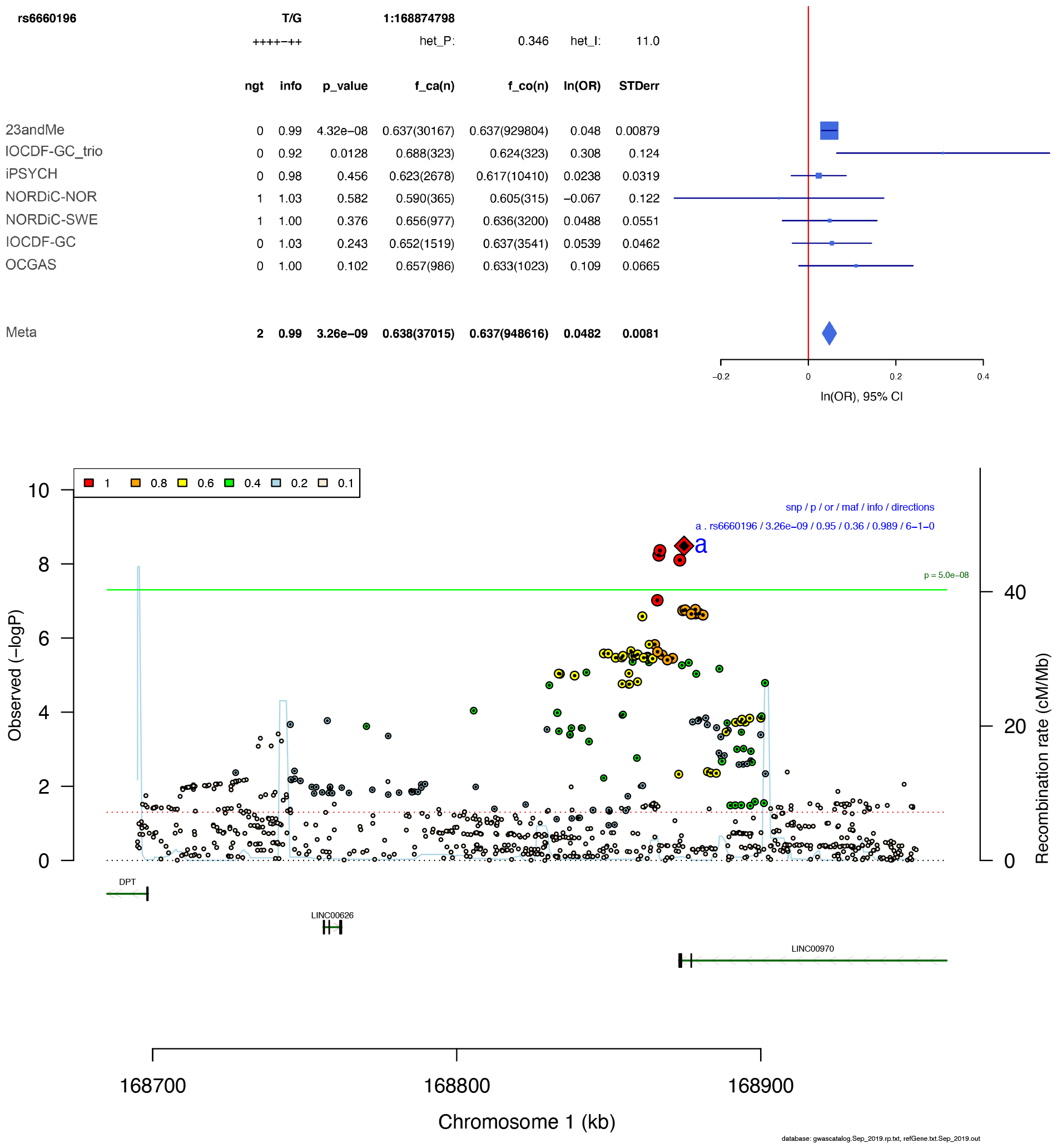
Forest plot (top) and regional association plot (bottom) of SNP rs6660196. Forest plot: Shows the imputation score (info), p-value (p_value), A1 allele frequency in cases and number of cases (f_ca(n)), A1 allele frequency in controls and number of controls (f_co(n)), effect of the association (ln(OR)), and standard error of the effect (STDerr) for each individual dataset and the over-all meta-analysis. On the right side, effects (ln(OR)) and 95% confidence intervals (CI) are plotted. At the top, the direction of effect for each study is shown (+ for positive effect of A1, - for negative effect of A1), and results of a test of heterogeneity (het_P and het_I) of effect across the individual datasets re displayed. Regional association plot: The –log10(P) of SNPs in the OCD meta-analysis GWAS is shown on the left y axis. The recombination rates expressed in centimorgans (cM) per Mb (Megabase) (blue line) are shown on the right y axis. Position in Mb is on the x axis. Only the SNPs with association p-value less than 0.1 were plotted. The lead SNP in the region is shown as a red diamond. Colour coding indicates LD to the lead SNP.

**Fig. S9.**
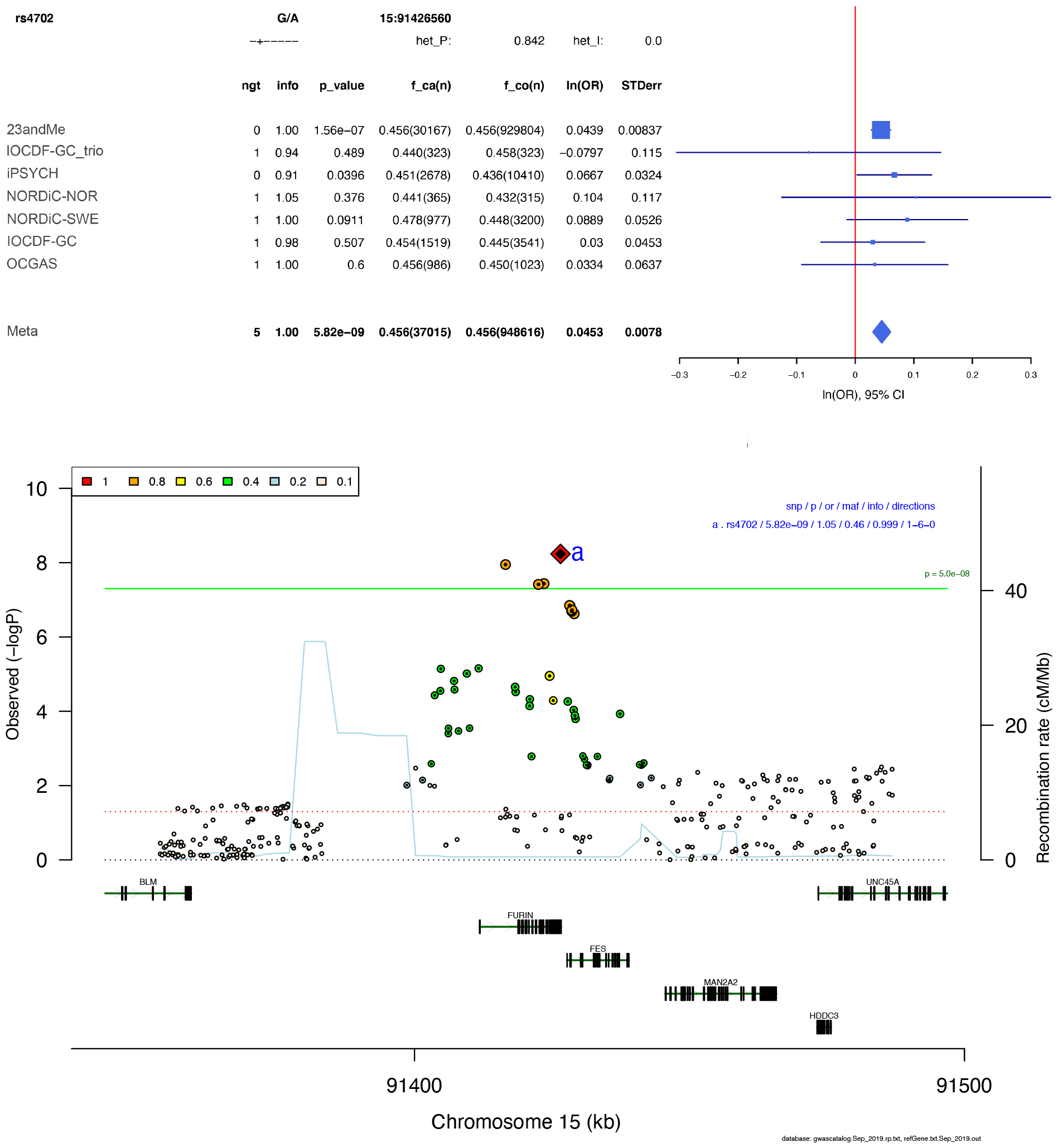
Forest plot (top) and regional association plot (bottom) of SNP rs4702. Forest plot: Shows the imputation score (info), p-value (p_value), A1 allele frequency in cases and number of cases (f_ca(n)), A1 allele frequency in controls and number of controls (f_co(n)), effect of the association (ln(OR)), and standard error of the effect (STDerr) for each individual dataset and the over-all meta-analysis. On the right side, effects (ln(OR)) and 95% confidence intervals (CI) are plotted. At the top, the direction of effect for each study is shown (+ for positive effect of A1, - for negative effect of A1), and results of a test of heterogeneity (het_P and het_I) of effect across the individual datasets re displayed. Regional association plot: The –log10(P) of SNPs in the OCD meta-analysis GWAS is shown on the left y axis. The recombination rates expressed in centimorgans (cM) per Mb (Megabase) (blue line) are shown on the right y axis. Position in Mb is on the x axis. Only the SNPs with association p-value less than 0.1 were plotted. The lead SNP in the region is shown as a red diamond. Colour coding indicates LD to the lead SNP.

**Fig. S10.**
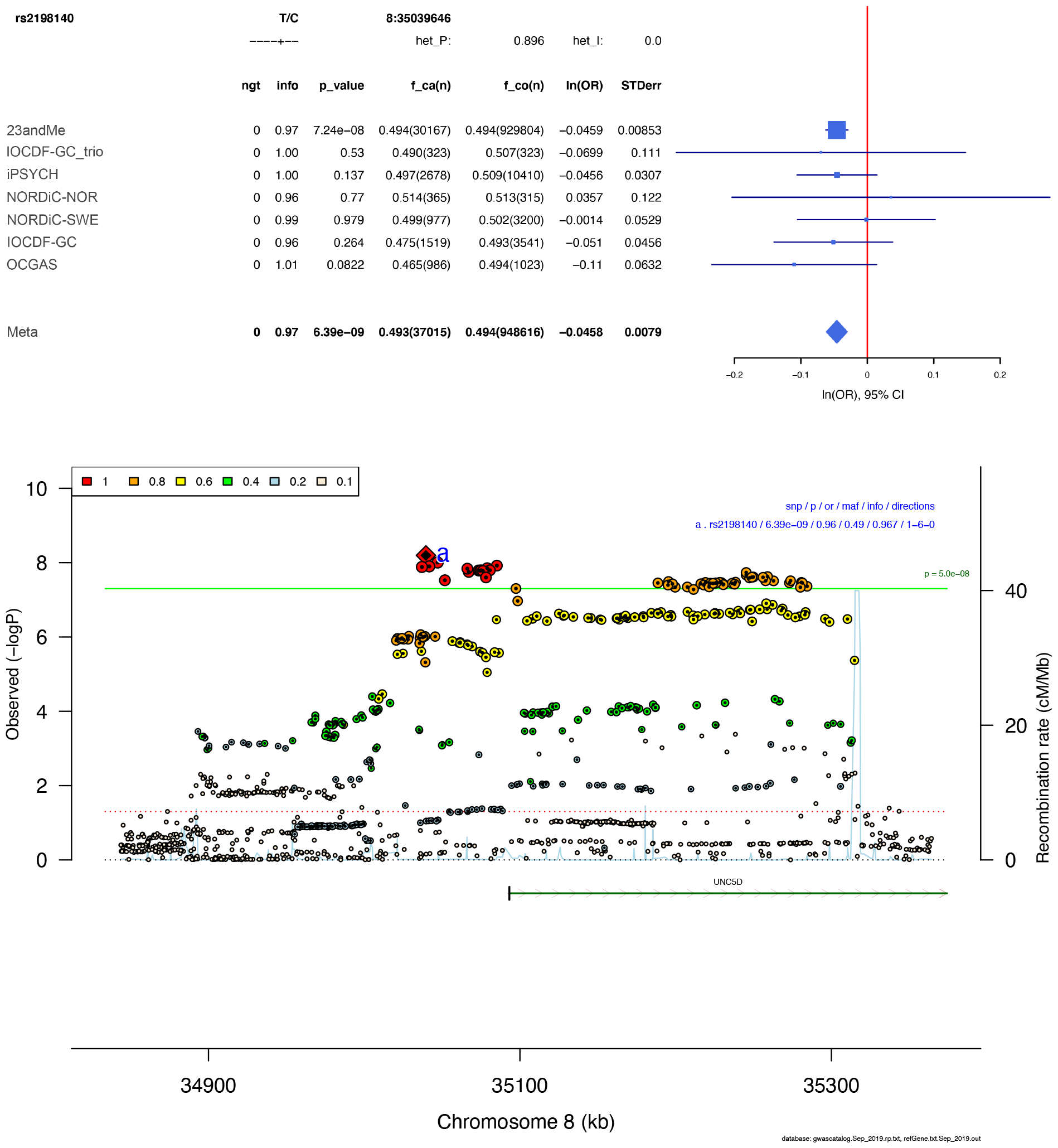
Forest plot (top) and regional association plot (bottom) of SNP rs2198140. Forest plot: Shows the imputation score (info), p-value (p_value), A1 allele frequency in cases and number of cases (f_ca(n)), A1 allele frequency in controls and number of controls (f_co(n)), effect of the association (ln(OR)), and standard error of the effect (STDerr) for each individual dataset and the over-all meta-analysis. On the right side, effects (ln(OR)) and 95% confidence intervals (CI) are plotted. At the top, the direction of effect for each study is shown (+ for positive effect of A1, - for negative effect of A1), and results of a test of heterogeneity (het_P and het_I) of effect across the individual datasets re displayed. Regional association plot: The –log10(P) of SNPs in the OCD meta-analysis GWAS is shown on the left y axis. The recombination rates expressed in centimorgans (cM) per Mb (Megabase) (blue line) are shown on the right y axis. Position in Mb is on the x axis. Only the SNPs with association p-value less than 0.1 were plotted. The lead SNP in the region is shown as a red diamond. Colour coding indicates LD to the lead SNP.

**Fig. S11.**
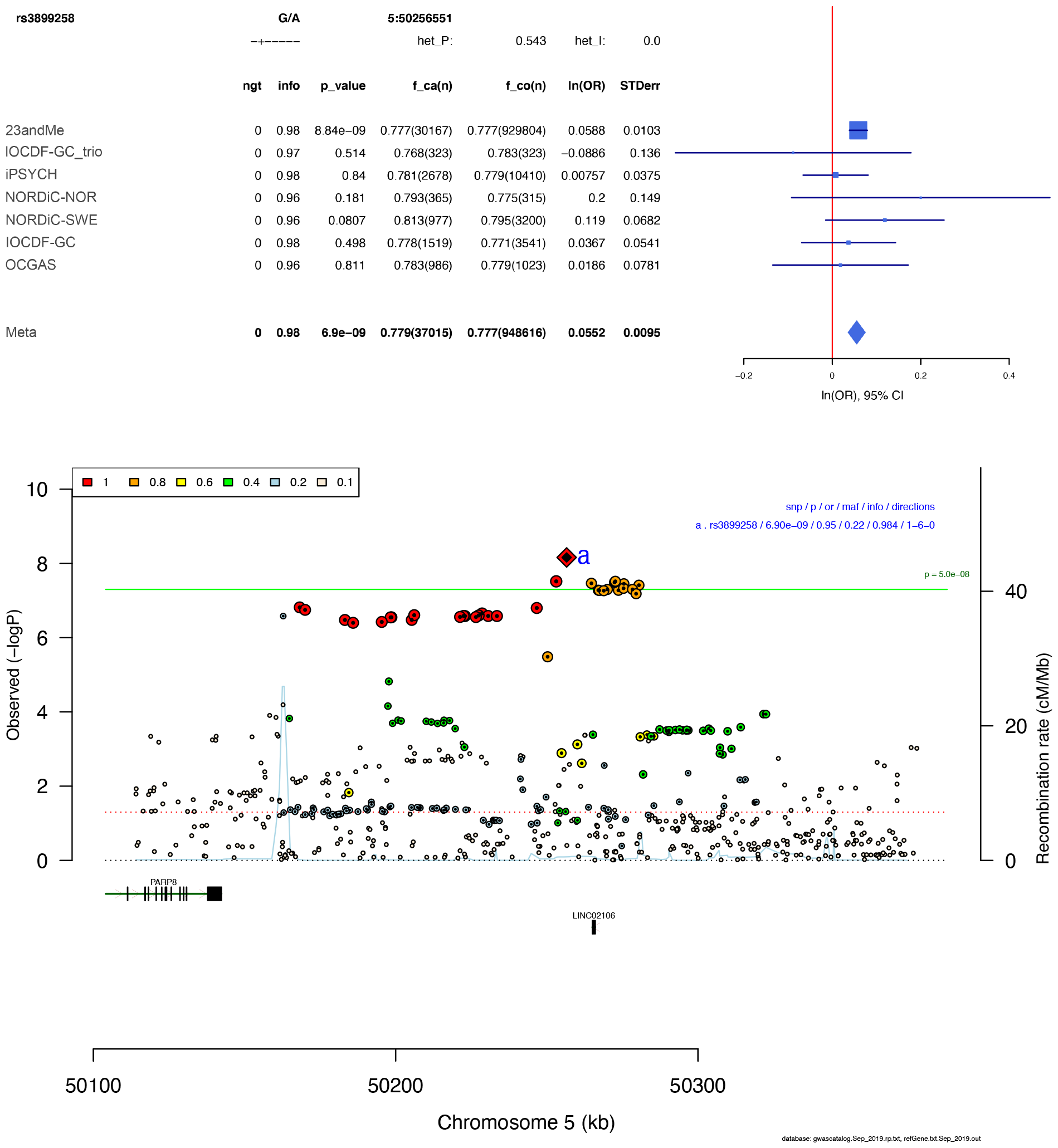
Forest plot (top) and regional association plot (bottom) of SNP rs3899258. Forest plot: Shows the imputation score (info), p-value (p_value), A1 allele frequency in cases and number of cases (f_ca(n)), A1 allele frequency in controls and number of controls (f_co(n)), effect of the association (ln(OR)), and standard error of the effect (STDerr) for each individual dataset and the over-all meta-analysis. On the right side, effects (ln(OR)) and 95% confidence intervals (CI) are plotted. At the top, the direction of effect for each study is shown (+ for positive effect of A1, - for negative effect of A1), and results of a test of heterogeneity (het_P and het_I) of effect across the individual datasets re displayed. Regional association plot: The –log10(P) of SNPs in the OCD meta-analysis GWAS is shown on the left y axis. The recombination rates expressed in centimorgans (cM) per Mb (Megabase) (blue line) are shown on the right y axis. Position in Mb is on the x axis. Only the SNPs with association p-value less than 0.1 were plotted. The lead SNP in the region is shown as a red diamond. Colour coding indicates LD to the lead SNP.

**Fig. S12.**
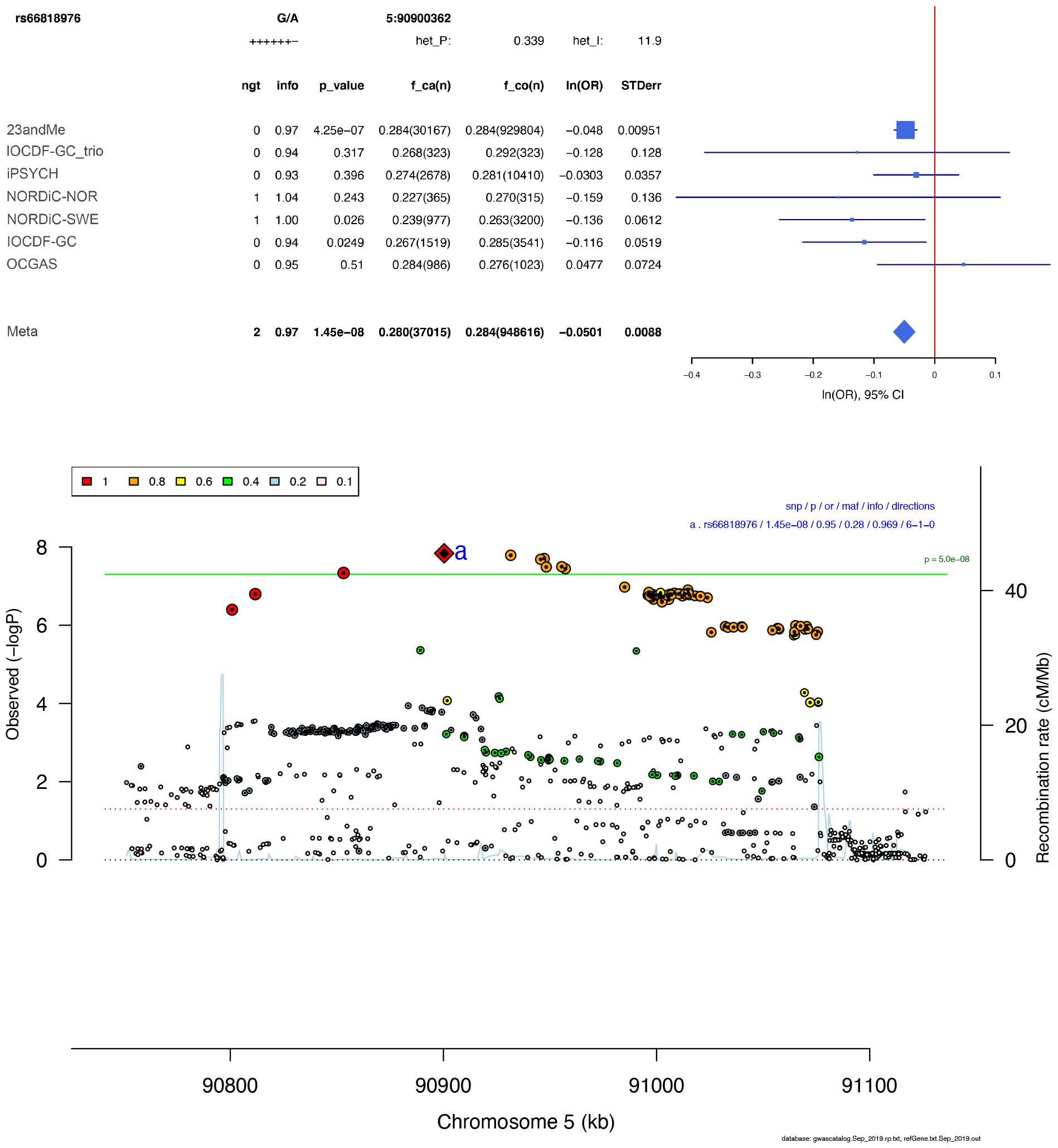
Forest plot (top) and regional association plot (bottom) of SNP rs66818976. Forest plot: Shows the imputation score (info), p-value (p_value), A1 allele frequency in cases and number of cases (f_ca(n)), A1 allele frequency in controls and number of controls (f_co(n)), effect of the association (ln(OR)), and standard error of the effect (STDerr) for each individual dataset and the over-all meta-analysis. On the right side, effects (ln(OR)) and 95% confidence intervals (CI) are plotted. At the top, the direction of effect for each study is shown (+ for positive effect of A1, - for negative effect of A1), and results of a test of heterogeneity (het_P and het_I) of effect across the individual datasets re displayed. Regional association plot: The –log10(P) of SNPs in the OCD meta-analysis GWAS is shown on the left y axis. The recombination rates expressed in centimorgans (cM) per Mb (Megabase) (blue line) are shown on the right y axis. Position in Mb is on the x axis. Only the SNPs with association p-value less than 0.1 were plotted. The lead SNP in the region is shown as a red diamond. Colour coding indicates LD to the lead SNP.

**Fig. S13.**
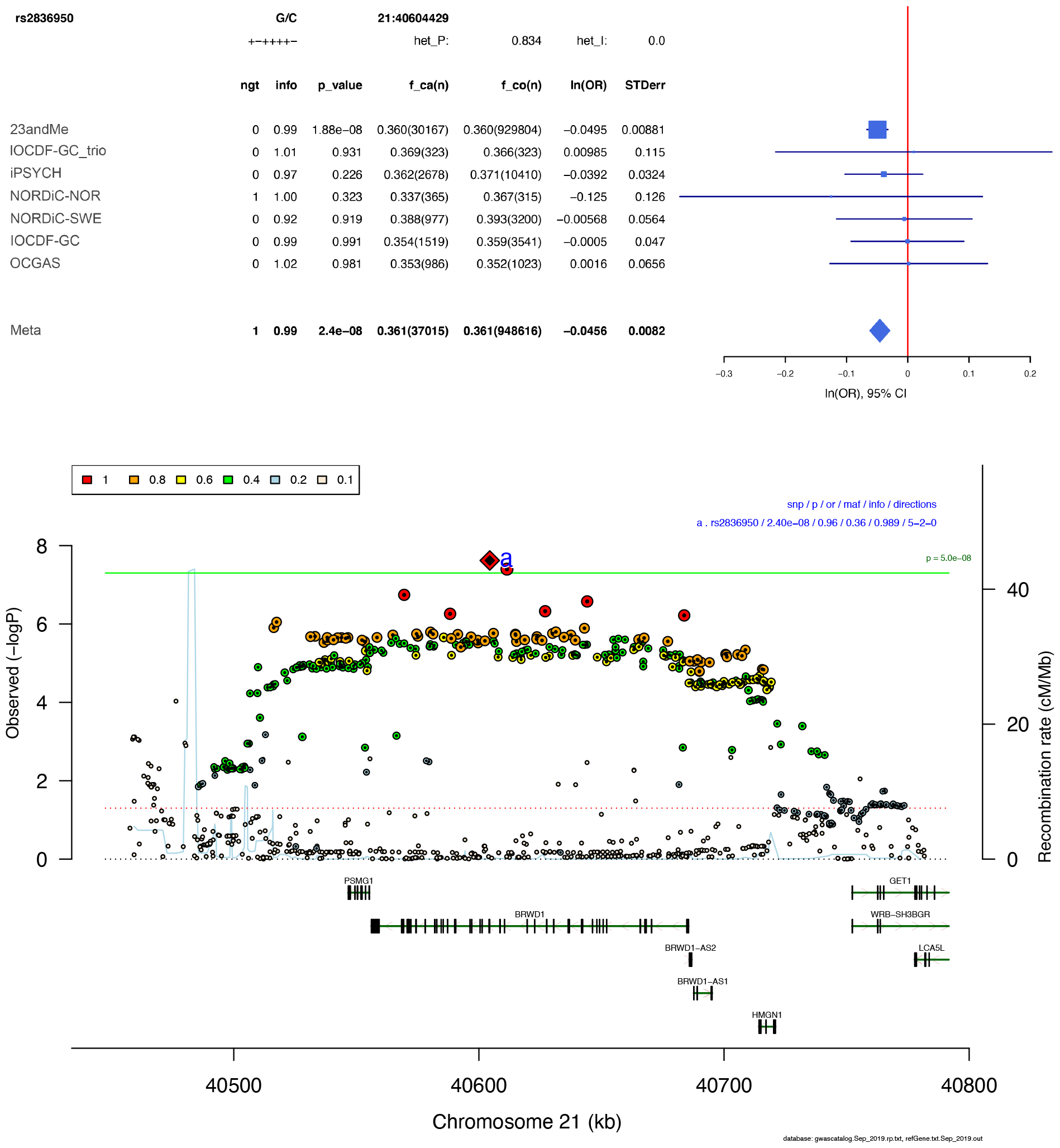
Forest plot (top) and regional association plot (bottom) of SNP rs2836950. Forest plot: Shows the imputation score (info), p-value (p_value), A1 allele frequency in cases and number of cases (f_ca(n)), A1 allele frequency in controls and number of controls (f_co(n)), effect of the association (ln(OR)), and standard error of the effect (STDerr) for each individual dataset and the over-all meta-analysis. On the right side, effects (ln(OR)) and 95% confidence intervals (CI) are plotted. At the top, the direction of effect for each study is shown (+ for positive effect of A1, - for negative effect of A1), and results of a test of heterogeneity (het_P and het_I) of effect across the individual datasets re displayed. Regional association plot: The –log10(P) of SNPs in the OCD meta-analysis GWAS is shown on the left y axis. The recombination rates expressed in centimorgans (cM) per Mb (Megabase) (blue line) are shown on the right y axis. Position in Mb is on the x axis. Only the SNPs with association p-value less than 0.1 were plotted. The lead SNP in the region is shown as a red diamond. Colour coding indicates LD to the lead SNP.

**Fig. S14.**
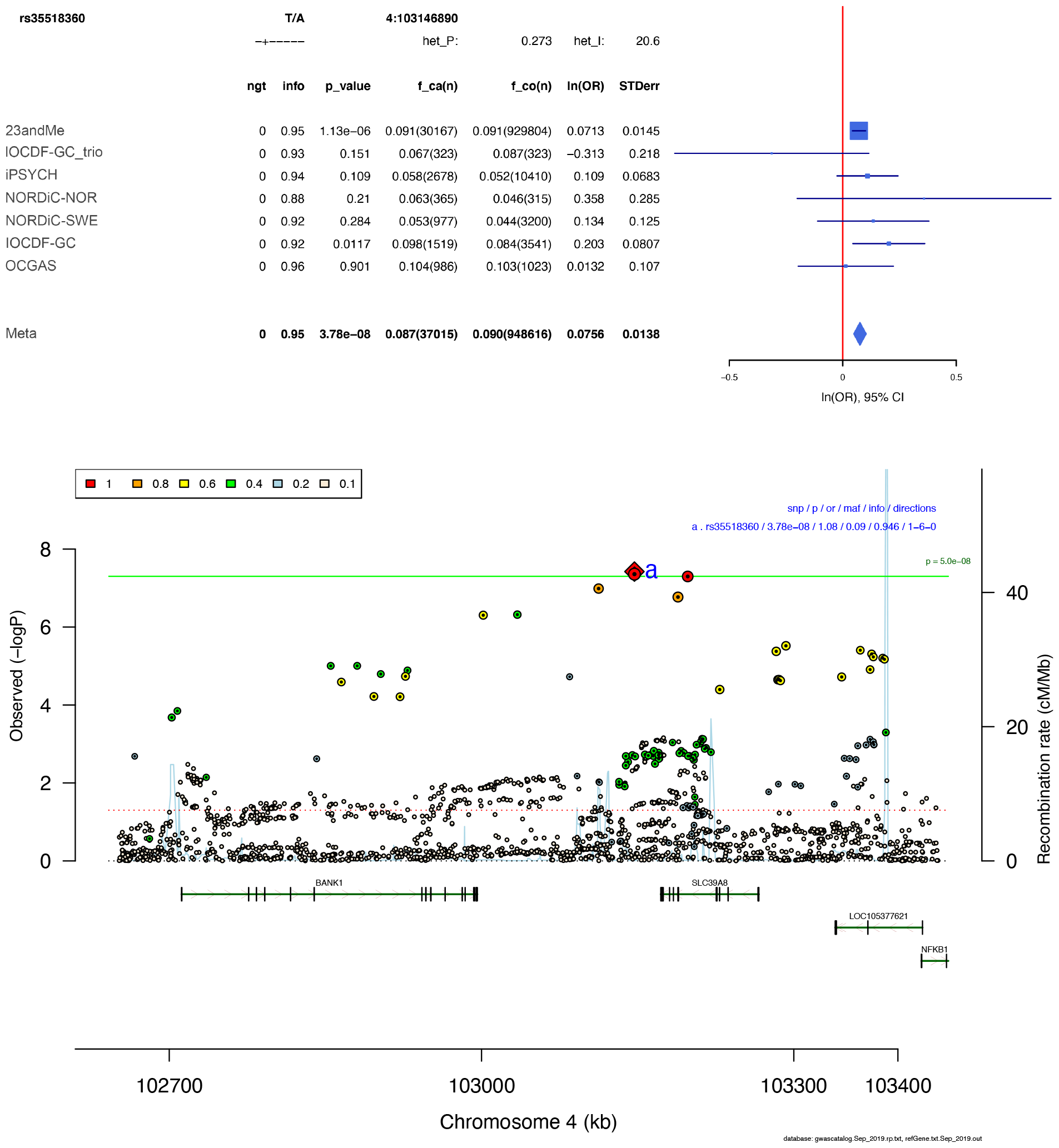
Forest plot (top) and regional association plot (bottom) of SNP rs35518360. Forest plot: Shows the imputation score (info), p-value (p_value), A1 allele frequency in cases and number of cases (f_ca(n)), A1 allele frequency in controls and number of controls (f_co(n)), effect of the association (ln(OR)), and standard error of the effect (STDerr) for each individual dataset and the over-all meta-analysis. On the right side, effects (ln(OR)) and 95% confidence intervals (CI) are plotted. At the top, the direction of effect for each study is shown (+ for positive effect of A1, - for negative effect of A1), and results of a test of heterogeneity (het_P and het_I) of effect across the individual datasets re displayed. Regional association plot: The –log10(P) of SNPs in the OCD meta-analysis GWAS is shown on the left y axis. The recombination rates expressed in centimorgans (cM) per Mb (Megabase) (blue line) are shown on the right y axis. Position in Mb is on the x axis. Only the SNPs with association p-value less than 0.1 were plotted. The lead SNP in the region is shown as a red diamond. Colour coding indicates LD to the lead SNP.

**Fig. S15.**
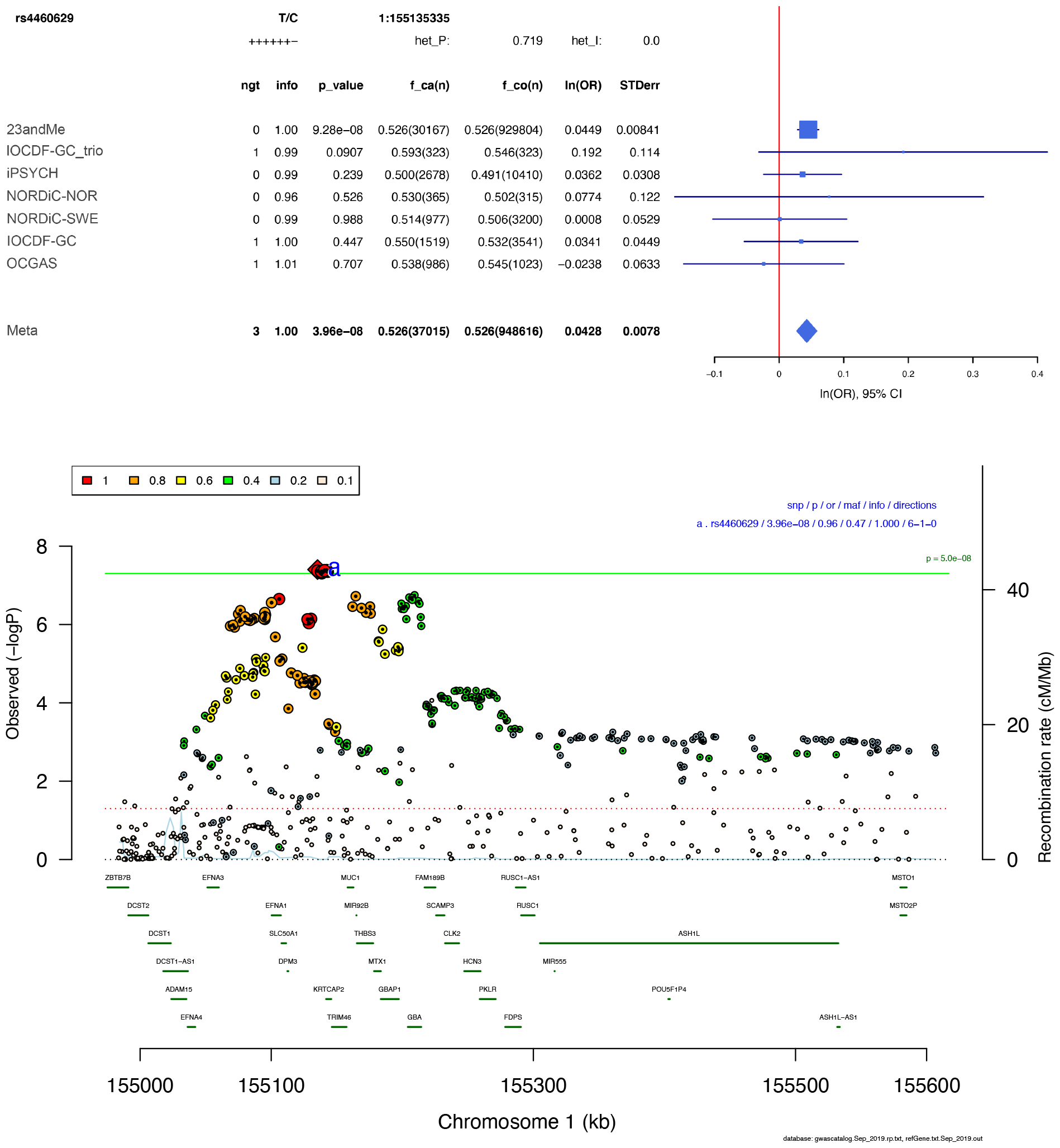
Forest plot (top) and regional association plot (bottom) of SNP rs4460629. Forest plot: Shows the imputation score (info), p-value (p_value), A1 allele frequency in cases and number of cases (f_ca(n)), A1 allele frequency in controls and number of controls (f_co(n)), effect of the association (ln(OR)), and standard error of the effect (STDerr) for each individual dataset and the over-all meta-analysis. On the right side, effects (ln(OR)) and 95% confidence intervals (CI) are plotted. At the top, the direction of effect for each study is shown (+ for positive effect of A1, - for negative effect of A1), and results of a test of heterogeneity (het_P and het_I) of effect across the individual datasets re displayed. Regional association plot: The –log10(P) of SNPs in the OCD meta-analysis GWAS is shown on the left y axis. The recombination rates expressed in centimorgans (cM) per Mb (Megabase) (blue line) are shown on the right y axis. Position in Mb is on the x axis. Only the SNPs with association p-value less than 0.1 were plotted. The lead SNP in the region is shown as a red diamond. Colour coding indicates LD to the lead SNP.

**Fig. S16.**
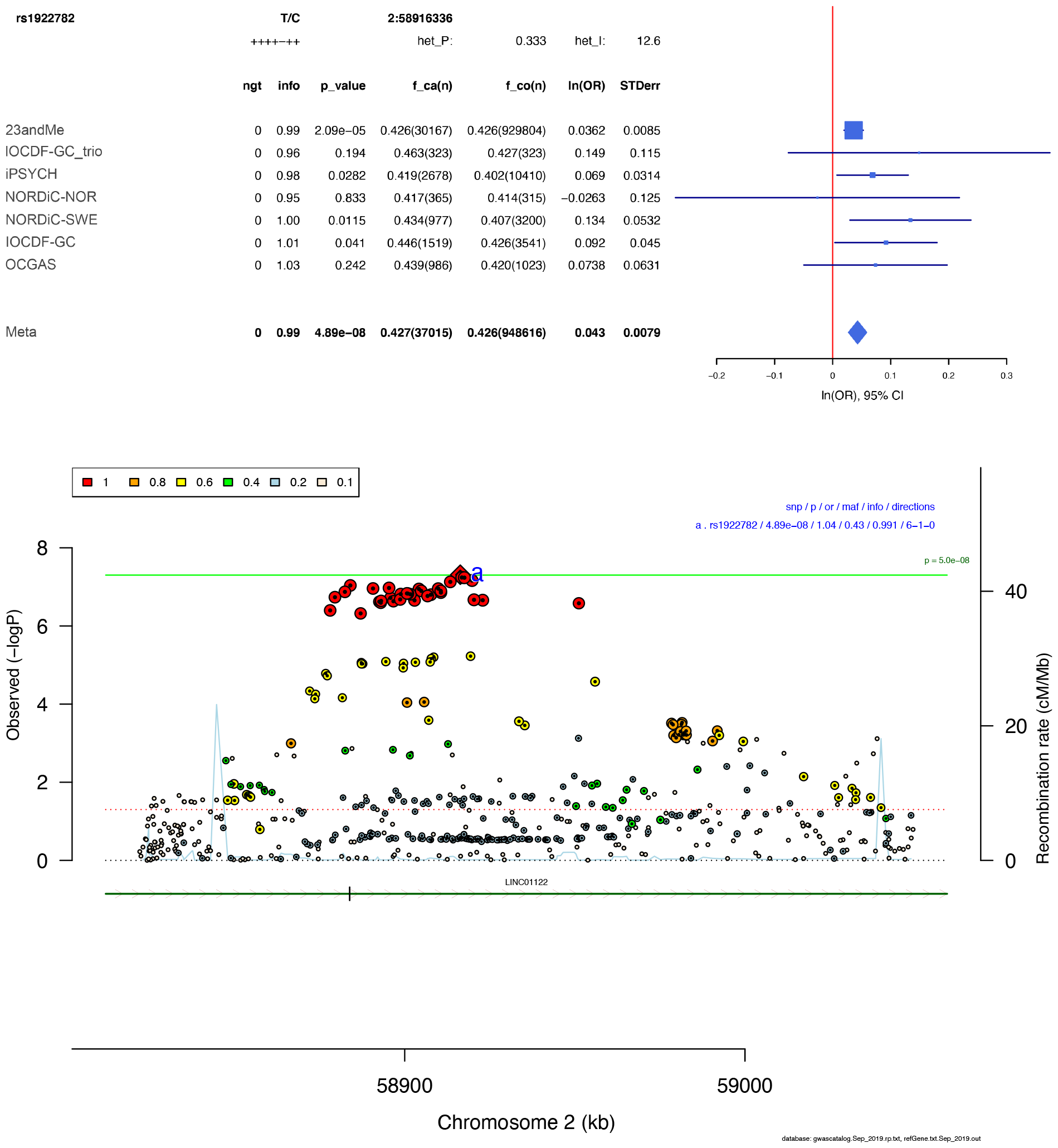
Forest plot (top) and regional association plot (bottom) of SNP rs1922782. Forest plot: Shows the imputation score (info), p-value (p_value), A1 allele frequency in cases and number of cases (f_ca(n)), A1 allele frequency in controls and number of controls (f_co(n)), effect of the association (ln(OR)), and standard error of the effect (STDerr) for each individual dataset and the over-all meta-analysis. On the right side, effects (ln(OR)) and 95% confidence intervals (CI) are plotted. At the top, the direction of effect for each study is shown (+ for positive effect of A1, - for negative effect of A1), and results of a test of heterogeneity (het_P and het_I) of effect across the individual datasets re displayed. Regional association plot: The –log10(P) of SNPs in the OCD meta-analysis GWAS is shown on the left y axis. The recombination rates expressed in centimorgans (cM) per Mb (Megabase) (blue line) are shown on the right y axis. Position in Mb is on the x axis. Only the SNPs with association p-value less than 0.1 were plotted. The lead SNP in the region is shown as a red diamond. Colour coding indicates LD to the lead SNP.

**Fig. S17.**
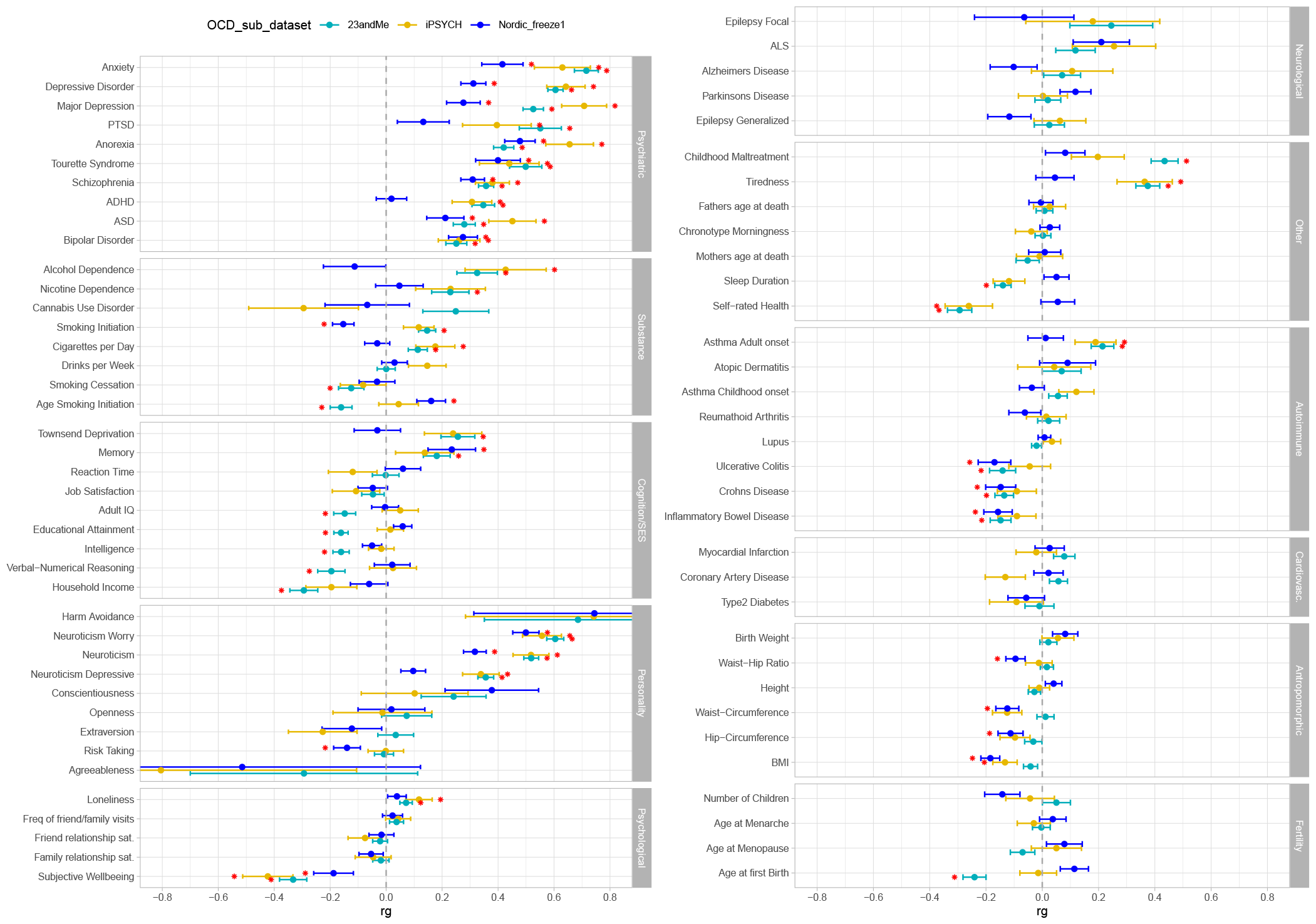
Genetic correlations (rg) between OCD, divided into three sub-groups, and behavioral, cognitive, psychiatric, neurologic, immunologic, metabolic, and anthropomorphic phenotypes. Error bars represent standard errors and red asterisks indicate significant associations after FDR correction for multiple testing.

**Fig. S18.**
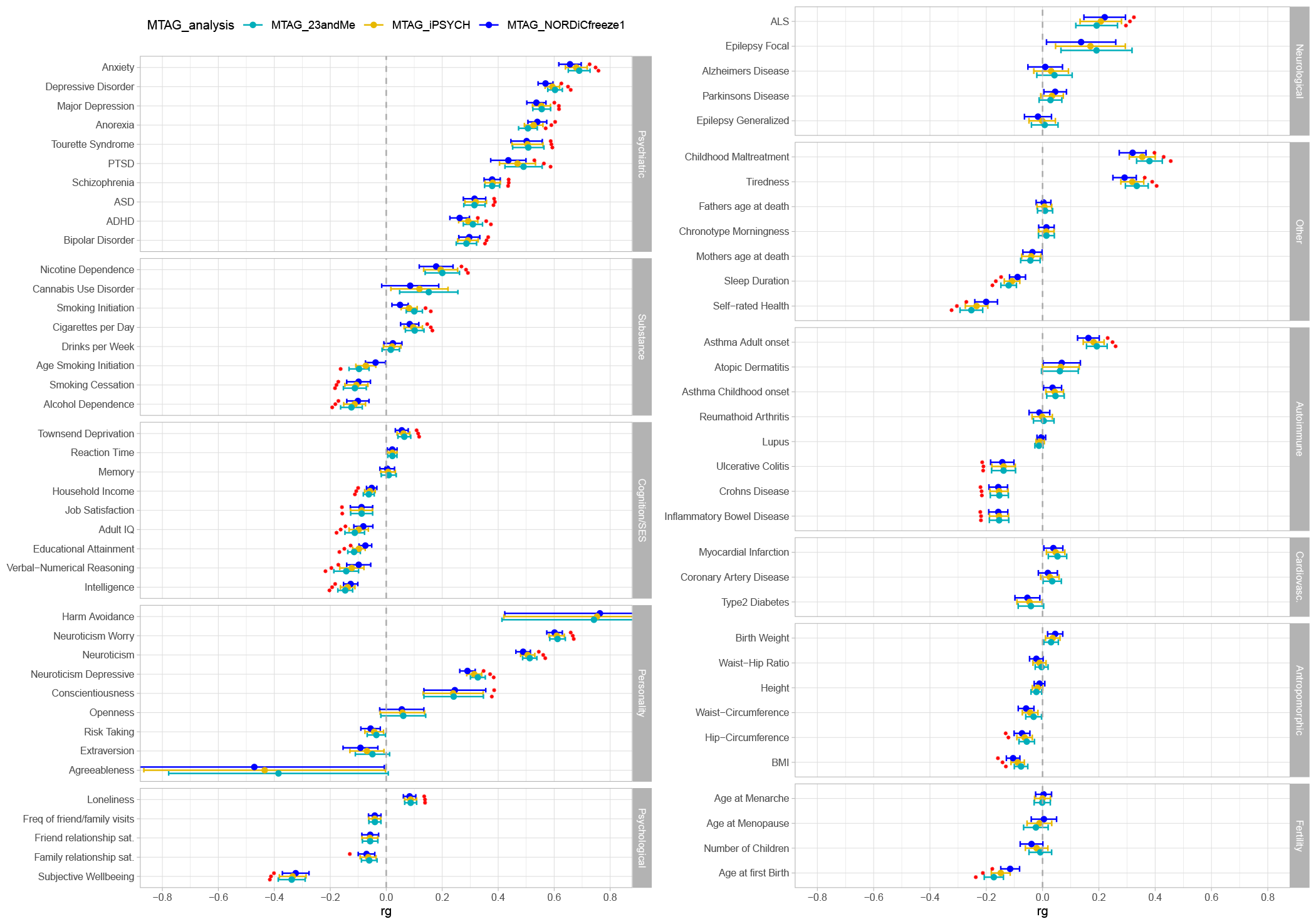
Genetic correlations (rg) between three MTAG analyses of OCD, and behavioral, cognitive, psychiatric, neurologic, immunologic, metabolic, and anthropomorphic phenotypes. Error bars represent standard errors and red asterisks indicate significant associations after FDR correction for multiple testing.

